# Oral Health Research Across the Lifespan: A Systematic Mapping Review of Cohort Studies in Australia and New Zealand

**DOI:** 10.1101/2025.07.17.25331745

**Authors:** Parsa Pirooz, Navodya Selvaratnam, Narendar Manohar, Mariam Al Asaad, Amit Arora

**Author notes:** Funding sources: N/A. Conflicts of interest: N/A.

## Abstract

**Objectives:** Oral health plays a crucial role in maintaining overall health and well-being, affecting over one-fifth of the population in Australia and New Zealand. Numerous cohort studies utilise The Life Course Health Development (LCHD) framework to explore oral health, highlighting the complex relationships among various influencing factors. However, a comprehensive synthesis of evidence on oral health-related cohort studies across the lifespan in Australia and New Zealand, which have unique demographic and environmental characteristics, is necessary to enhance the understanding of the diverse existing research in these regions. Therefore, this systematic mapping review aims to identify oral health-related cohort studies in Australia and New Zealand, providing details on their demographics, methods, oral health measurements, and a framework of the investigated outcomes and explanatory factors.

**Methods:** A systematic search was conducted across five electronic databases: MEDLINE (OVID), Embase (OVID), Web of Science (ISI), Scopus, and CINAHL followed by backward citation chasing. Oral health-related cohort studies conducted in Australia and New Zealand were included. A descriptive synthesis approach was employed to summarise the studies, and results were reported in accordance with PRISMA-ScR guidelines.

**Results:** From the 226 included publications, a total of 37 cohort studies involving primary data collection and ten independent data linkage studies were identified. The geographical distribution, general characteristics, and design aspects of the identified studies were summarised. Seventy oral health measurements employed in these studies were identified and categorised. Additionally, an Evidence and Gap Map (EGM) was presented to illustrate links between 38 themes for explanatory factors and 32 themes for outcomes in the identified studies.

**Conclusion:** Numerous oral health-related cohort studies in Australia and New Zealand reveal significant variations. Over-represented study populations and measurements provide opportunities for secondary research, while under-represented areas should be the focus of future primary research. This review serves as a roadmap for researchers, policymakers, and clinicians in making evidence-based decisions to improve oral health.

## 1. Introduction

Oral health is an integral component of overall health and well-being, as it has a multifaceted influence on the physical, psychological, emotional, and social aspects of health, thereby influencing overall quality of life (1, 2). Oral health conditions, including dental caries, periodontitis, and edentulism, rank among the most common health issues, impacting 3.69 billion people worldwide in 2021 (3). High-income countries such as Australia and New Zealand also face the burden of oral health conditions. In Australia, poor oral health contributed to 4.5% of all the burden caused by non-fatal diseases in 2022 (4). In 2017-18, approximately 26% of children aged 5 to 14, 33% of adults aged 15 to 64, and 27% of those over 65 were reported to have at least one tooth with untreated dental caries (5), while 29% of adults aged 15 and over experienced gingivitis (4) in Australia. Similarly, around one-fifth of the New Zealand population have been reported to suffer from periodontal disease and dental decay in 2019 (6). In a study in 2023, the average decayed, missing, or filled primary teeth (dmft) scores were calculated as 1.95 for five-year-old children and 0.74 for children aged 12-13 years, respectively (7). Hence, addressing these oral health issues is vital for improving the overall health outcomes of populations in both Australia and New Zealand.

In recent years, the Life Course Health Development (LCHD) framework approach to oral health research has gained attention. It explains health as the result of the interplay among various genetic, biological, behavioural, social, and economic factors throughout human life (8). Adopting the LCHD framework provides a powerful approach to understanding how oral diseases develop, shaped by oral health factors, associated inequalities, and their interconnections with overall health (9). This is particularly relevant given that the most common oral health conditions, including those previously mentioned, are chronic and arise from prolonged exposure to their corresponding risk/protective (explanatory) factors (10, 11). The LCHD’s comprehensive view on health is enabled by the cohort study design, as it allows for establishing connections between exposures and outcomes (either prospectively or retrospectively) over an extended temporal framework (12, 13), while creating an opportunity to concurrently assess oral health and other general health domains in a single study (14, 15).

Numerous cohort studies with a primary or secondary focus on oral health have been conducted globally. In 2019, a research workshop in Bangkok resulted in a scoping review that mapped and detailed the characteristics of global oral health-related birth cohort studies, serving as a preliminary step towards establishing a global consortium dedicated to these cohorts (GLOBICS) (16). However, focusing solely on birth-cohort studies excludes a significant number of cohort studies conducted at other life stages. Research on older cohorts is crucial for understanding oral health, as significant conditions like periodontal diseases, edentulism, and oral cancers often arise later in life rather than in early stages (17–19). Moreover, GLOBICS publications (16, 20–22) had a global reach rather than a regional focus, which may have resulted in a loss of details pertinent to specific demographic regions. For example, the details of exposure and outcome from just four cohort studies in Australia and New Zealand were presented in the published review (20). Therefore, a more focused regional analysis is required for a better understanding of life course oral health research in the context of Australia and New Zealand.

Australia and New Zealand are home to distinctly diverse populations, comprising Indigenous communities—including Aboriginal and Torres Strait Islander peoples in Australia and Māori and Pacific Islanders in New Zealand—as well as a variety of migrant communities from Europe, Asia, the Middle East, and Africa, all of which significantly influence health practices and outcomes across the life course (23, 24). One of the unique health issues is the poorer oral health outcomes of Indigenous populations, compared to the general population, which is well-established within the literature (25–27). As a result, the investigation into this health disparity has driven a few previously conducted cohort studies in the region, namely the Aboriginal Birth Cohort (ABC) study in Australia and the Pacific Islands Families (PIF) study in New Zealand (28, 29). Aside from the demographic factors of health in this region, variations in other social determinants of health including, but not limited to, higher income, reduced inequality, improved education levels, and comprehensive health policies as well as controlled environmental factors (i.e., fluoridation of community water supplies) would impact oral health outcomes, thus emphasising the necessity for a specific investigation into oral health in Australia and New Zealand (30–33).

Australia and New Zealand have hosted several cohort studies that collected oral health data. In New Zealand, the Dunedin Multidisciplinary Health and Development Study (DMHDS) is recognised as one of the most enduring cohort studies globally, spanning more than five decades and focusing on oral and various general health issues (34). Similarly, Australia’s “45 and Up Study” represents another large-scale multidisciplinary cohort study that recruited individuals aged 45 and over at the baseline, following them for more than 15 years now (35). Additionally, other cohort studies, such as the Health Smiles Healthy Kids (HSHK) study and the Study of Mothers’ and Infants’ Life Events Affecting Oral Health (SMILE), were specifically designed and executed to understand and address socioeconomic inequality in oral health (36, 37). These examples offer only a glimpse into the landscape of oral health-related cohort studies in Australia and New Zealand. However, no synthesis of evidence has been performed on oral health-related cohort studies across the lifespan in this region to capture the complete picture.

Systematic mapping reviews, along with the Evidence and Gap Map (EGM) as a visual synthesis tool that illustrates the concentration of available evidence, aim to broadly identify, describe, and catalogue existing literature on a research topic (38, 39). This is in contrast to traditional systematic reviews, which focus on synthesising evidence from primary research to answer specific questions (39). Applying this methodology to the landscape of life course oral health in Australia and New Zealand provides several benefits for researchers, policymakers, and other interested stakeholders. It will outline and detail the existing literature, aiding in the identification of well-established research areas and evidence gaps within the field (40). Identifying heavily researched areas highlight valuable resources for potential secondary analysis (i.e. systematic reviews, pooled analysis, and data linkage) and inform evidence-based policy decisions and targeted interventions. Meanwhile, detected gaps in the literature can help set priorities for future primary research, funding decisions, and strategic resource allocation to address under-investigated fields and populations (40–42). Therefore, this systematic mapping review aims to identify oral health-related cohort studies across Australia and New Zealand, summarising their demographic and methodological characteristics, cataloguing the employed oral health measurements, and presenting an EGM of the explored explanatory factors and outcomes from these studies.

## 2. Methods

This systematic mapping review follows the reporting guidelines outlined in the Preferred Reporting Items for Systematic Reviews and Meta-Analyses extension for Scoping Reviews (PRISMA-ScR) Checklist (Appendix 1) (43). The protocol of this systematic review has been registered with the Open Science Framework (OSF) (osf.io/2948n) (44, 45).

### 2.1 Eligibility Criteria

This review used the Population-Concept-Context (PCC) framework, with the addition of the study design (S), to inform its eligibility criteria (46) (see Table 1).

**Table 1.** Eligibility criteria according to the PCC framework.

|  | Inclusion Criteria | Exclusion Criteria |
| --- | --- | --- |
| **Population** | • Children and adults across the lifespan<br>• Any gender<br>• Any ethnicity, culture, or race<br>• Any socio-economic status | • Animal models or in vitro research |
| **Concept** | • Oral and dental health-related measurements collected within cohort studies<br>• Self-reported or clinically measured data |  |
| **Context** | • Studies conducted primarily within Australia and/or New Zealand<br>• National, state-level, or specific regional cohorts based in either or both countries |  |
| **Study Design** | • Reporting on prospective or retrospective cohort studies with or without data linkage | • Commentary articles<br>• Poster and Abstracts<br>• Randomized Controlled Trials, Quasi-experiments<br>• Systematic reviews, literature reviews, mini-reviews<br>• Qualitative Studies<br>• Book Chapters |

### 2.2 Information Sources

Five electronic databases were systematically searched: MEDLINE (OVID), Embase (OVID), Web of Science (ISI), Scopus, and CINAHL. Additionally, backward citation chasing (i.e., searching reference lists manually) was conducted on the included studies to ensure an extensive literature search. The initial search was conducted on 20 December 2023 and will be updated in July 2025 prior to journal submission.

### 2.3 Search Strategy

Review questions and related search terms were devised based on the eligibility criteria outlined by the PCC framework. A set of predetermined keywords, medical subject headings (MESH) and the Boolean operators (‘AND’ and ‘OR’) was used to run a test search in MEDLINE (OVID), subsequently adapting it to other four databases (see Appendix 2). This process was carried out in collaboration with an experienced librarian in health sciences (K.E.). No restrictions on the publication date, language, study type, or region were placed to ensure that all relevant studies were captured in the initial electronic database search.

### 2.4 Screening and Study Selection Process

Identified records were imported into the systematic review tool Covidence^©^ (Veritas Health Innovation Ltd, Australia), and duplicates were automatically removed. The study selection process involved two screening stages. Two reviewers (N.S. and P.P.) independently screened titles and abstracts against the eligibility criteria and then independently assessed the full texts of potentially relevant articles for final inclusion. For studies with uncertain eligibility, authors were contacted up to three times for clarification. If no response was received, their eligibility was evaluated based on existing information.

Reviewer disagreements were resolved by discussion with a third reviewer (A.A.). The reasons for exclusion at the full-text screening stage were documented (see Appendix 3). Furthermore, backward citation chasing was conducted to identify additional relevant publications. The study selection process was performed based on the Preferred Reporting Items for Systematic Reviews and Meta-Analyses extension for Scoping Reviews (PRISMA-ScR) checklist and presented as a flow diagram in the results section (43).

### 2.5 Data Extraction

A standardised data extraction form was developed inspired by the data extraction template for scoping review by Joanna Briggs Institute (JBI) Manual for Evidence Synthesis (46). The data extraction form was calibrated and pilot-tested with two studies to ensure reviewer consistency and comprehensive data extraction. Three reviewers (N.S., P.P., and M.A.) independently extracted the data from all included studies, which was subsequently confirmed by two other reviewers (A.A. and N.M.). The following information was extracted from each included study: cohort study name, study type, location, baseline date, eligibility criteria, age at recruitment, age at data collection waves, investigated variables, baseline sample size, ethnicity at baseline, and included intervention (if any) (see Appendix 4).

### 2.6 Data Synthesis

A descriptive synthesis approach was employed to summarise the extracted data, focusing on mapping the extent, range, and nature of the studies rather than analysing specific study findings in depth. The key characteristics of the included cohort studies were presented in tables and bar charts. The geographical distribution of the cohort studies was plotted on a map using the Felt^©^ mapping tool (Felt LLC, USA). Oral health measurements were selected among the identified investigated variables for each study and presented using an interactive bipartite network. Additionally, the investigated variables were categorised and classified as explanatory factors or outcomes for each publication of the cohort studies, leading to the creation of an EGM. All figures, apart from the PRISMA flowchart and geographical map, were created using HTML codes. One reviewer (P.P.) initially performed the data synthesis process, which was subsequently verified by two other reviewers (A.A., N.M.).

## 3. Results

Initially, a total of 7,497 records were identified through all selected electronic databases (n = 7,460) and citation chasing (n = 37). After automatically and manually removing duplicates, 3,282 titles and abstracts were retrieved for further examination. Post review, 3,044 records were excluded, leading to 238 full-text publications being retrieved for reading. After excluding articles in the full-text reading stage (n = 12, see Appendix 3), a total of 226 publications met the eligibility criteria and were included. Based on these publications, 37 cohort studies with primary data collection and 10 independent data linkage studies, which relied solely on registries and records, were included in this systematic mapping review. The references for the included studies and their publications are classified and provided in Appendix 5. The identification, screening, and selecting stages is presented as a PRISMA flow diagram (See Figure 1).

**Figure 1.**
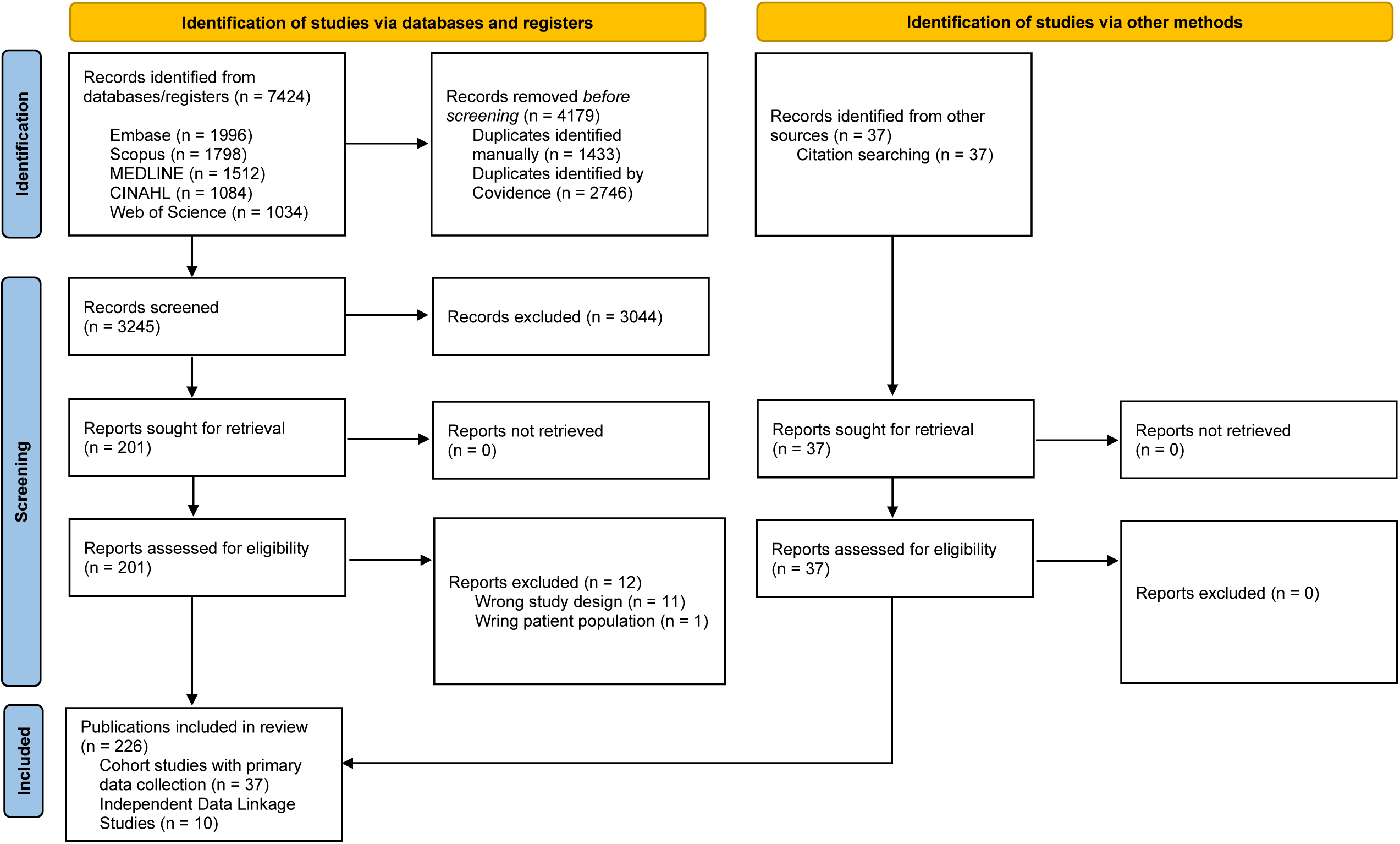
Preferred Reporting Items for Systematic Reviews and Meta-Analyses (PRISMA) flow diagram of literature search and study selection process.

Figure 2 shows the trend in publications of the included studies. The greatest number of publications was in 2020 (n = 17). While we found publications dating back to 1980, most of them (92.5%) were published from 2010 onwards (66.8%).

Figure 3 illustrates the geographical distribution of the identified studies across Australia and New Zealand, indicating that 37 studies were based in Australia and 10 in New Zealand. In Australia, there were five national studies and 32 state-based ones. In New Zealand, three studies were national, while seven were state-based. South Australia serves as the hub for most of the oral health-related cohort studies in the region, with a total of seven state-inclusive cohort studies.

**Figure 3.**
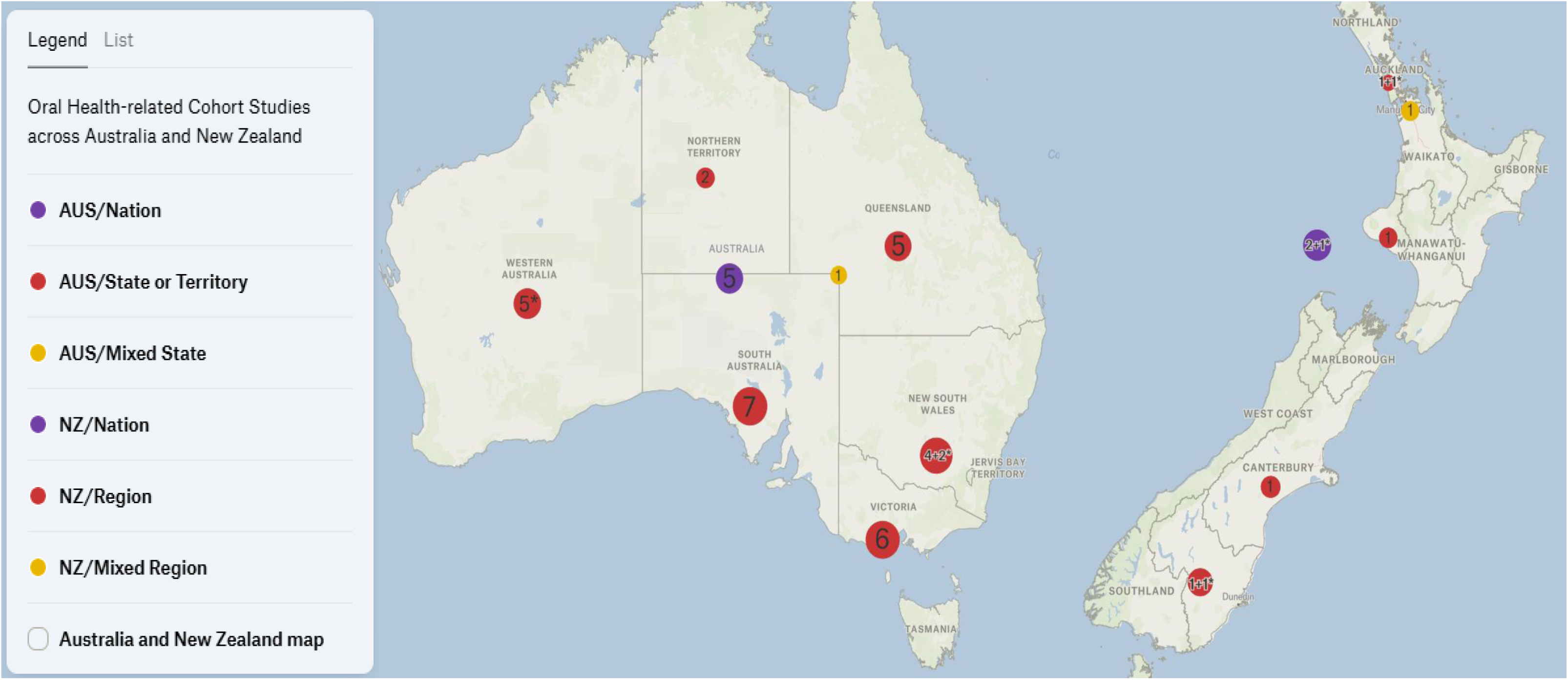
Geographical distribution of 47 cohort studies related to oral health across Australia and New Zealand, comprising 37 primary cohort studies and 10 independent data linkages^1^.

### 3.1 Cohort Studies with Primary Data Collection

The general characteristics of the primary cohort studies are summarised in Table 2. There are 14 studies that collected data in urban settings, five studies in rural areas, and 17 studies encompassing both settings. Amongst all included cohort studies, the earliest dated study with oral health-related data is the “University of Adelaide longitudinal study of growth and development” study, which has its baseline data collected over a 20-year period from 1950 to 1970. The DMHDS is the most prolific oral health-related cohort study in the region, resulting in the publication of 70 articles that present various aspects of oral health from this study.

**Table 2.** General characteristics of cohort studies with primary data collection presenting oral health data.

| Cohort study name (Abbreviation) | Australian State or Territory/New Zealand Region, Country | Setting<sup>2</sup> | Cohort baseline | Primary study population | N<sup>3</sup> |
| --- | --- | --- | --- | --- | --- |
| 1. Australian Research Centre for Population Oral Health Cohort Study—The University of Adelaide | South Australia, Australia | U, R | 1988-1989 | Children who have not previously undergone an orthodontic treatment, attending Dental Health Services (SDS) | 6 |
| 2. Mater Mother's Hospital Study_2003 | Queensland, Australia | U | 2003 | Pre-term (< 37weeks' gestation with birthweights < 2000 g) and full-term infants with normal birthweights (> 2500 g), recruited at birth from the Mater Mothers' Hospital | 2 |
| 3. Aboriginal Birth Cohort Study (ABC) | Northern Territory, Australia | U | 1987-1990 | Live-born singletons to a mother identifying as Aboriginal or Torres Strait Islander at the Royal Darwin Hospital | 9 |
| 4. University of Adelaide Twin Study (UAT) | Cohort 1: South Australia, Australia | U, R | 1983 - 1993 | Twins, recruited from the Australian Twin Registry, the Australian Multiple Birth Association, newspaper birth announcements, hospitals, and prenatal exercise classes | 19 |
|  | Cohort 2: South Australia and Victoria, Australia |  | 1995 |  |  |
|  | Cohort 3: Australia |  | 2005 |  |  |

<sup>2</sup> Urban: U, Rural: R<sup>3iii</sup> Number of published articles presenting oral health data. Please note that articles presenting data from more than one cohort study have been counted towards the total number of published articles for all of the respective cohort studies.

|  |  |  |  |  |  |
| --- | --- | --- | --- | --- | --- |
| **5.** Christchurch Child Development Study | Canterbury, New Zealand | U | 1977 | Children born in maternity units in the Christchurch urban region | 3 |
| **6.** Concord Health and Ageing in Men Project (CHAMP) | New South Wales, Australia | U | 2005 - 2006 | Men living in a defined geographical region (the Local Government Areas of Burwood, Canada Bay, and Strathfield), registered in the New South Wales Electoral Roll, and not living in a residential aged care facility | 8 |
| **7.** Dunedin Multidisciplinary Health and Development Study (DMHDS) | Otago, New Zealand | U | 1975-1976 | Children born at the Queen Mary Hospital, Dunedin | 70 |
| **8.** Environments for Healthy Living – the Griffith Birth Cohort study (EFHL) | Queensland, Australia | U, R | 2006-2011 | Births from the three public maternity hospitals in the participating districts (Logan, Gold Coast and The Tweed Hospitals) | 4 |
| **9.** Gudaga Study | New South Wales, Australia | R | 2005-2007 | Children with either of the biological parents identified as Aboriginal born in the Macarthur region | 3 |
| **10.** Longitudinal Study of Australian Children (LSAC) | Australia | U, R | 2004 | Children from urban and rural areas of all states and territories in Australia in the federal government's universal health insurance (Medicare) enrolment database identified by Indigenous indicator on their record | 12 |
| **11.** Longitudinal Study of Indigenous Children (LSIC) | Australia | U, R | 2008 | Children from remote, regional and urban locations of all states and territories in Australia | 5 |
| **12.** Pacific Islands Families Study (PIF) | Auckland, New Zealand | U | 2000 | Pacific infant singletons or first-borns in multiple births at Middlemore Hospital with at least one parent identified as being of Pacific Islands ethnicity and a New Zealand permanent resident | 5 |
| **13.** Queensland Birth Cohort Study | Queensland, Australia | R | 2007-2008 | Mother-child pairs attending public birthing and community health clinics of The Logan-Beaudesert district | 5 |
| **14.** South Australian Dental Longitudinal Study (SADLS) | South Australia, Australia | U, R | SADLS I:<br>1991-1992 | A stratified random sample of non-institutionalized people aged $\geq 60$ years, residing in two South Australian cities (Adelaide and Mount Gambier), selected from the South Australian State Electoral Database | 10 |
|  |  |  | SADLS II:<br>2013-2014 |  |  |
| **15.** Study of Mothers' and Infants' Life Events Affecting Oral Health (SMILE) | South Australia, Australia | U | 2013-2014 | Newborns at the main three public hospitals in Adelaide | 9 |
| **16.** The 45 and Up Study | New South Wales, Australia | U, R | 2006-2009 | Men and women residing in the State of New South Wales, registered in the Medicare Australia enrolment database | 6 |
| **17.** The Child Fluoride Study (CFS) | South Australia and Queensland, Australia | U, R | CFS Mark 1:<br>1991-1992 | A) Children enrolled in the school dental service (SDS) of South Australia | 4 |
|  |  |  |  | B) Children enrolled in the school dental service (SDS) of Townsville and Brisbane, Queensland |  |
|  |  |  | CFS Mark 2:<br>2002-2003 | Children enrolled in the school dental service (SDS) of South Australia |  |
| **18. Peri/Postnatal Epigenetic Twins Study (PETS)** | Victoria, Australia | U | 2007 | Mothers and their twins recruited from multiple pregnancy clinics at three Melbourne hospitals midway through their second trimester (18–22 weeks gestation) | 3 |
| **19. VicGeneration study (VicGen)** | Victoria, Australia | U, R | 2008 | Newborns residing in metropolitan (local government areas (LGAs) of Brimbank and Maribyrnong), regional (city of Ballarat), and rural (50-100km radius around Ballarat) areas of Victoria, without a complex medical condition (newborn), mental illness (parents), and an intention to relocate within the next 12 months | 7 |
| **20. Growing Up in New Zealand** | Auckland and Waikato, New Zealand | U, R | 2009-2010 | Pregnant women residing in the three District Health Boards (DHBs) of Auckland, Counties-Manukau, and Waikato | 1 |
| **21. Healthy Smiles Healthy Kids (HSHK)** | New South Wales, Australia | U | 2009-2010 | Mother-infant pairs in public hospitals located within the Sydney and South Western Sydney Local Health Districts (LHDs) | 2 |
| **22. South Australian Aboriginal Birth Cohort Study (SAABC)** | South Australia, Australia | U, R | 2011-2012 | Pregnant women with an Aboriginal child who were residing in South Australia, recruited through the antenatal clinics of South Australian Aboriginal Community Controlled Health Organisations and hospitals | 1 |
| **23. Splash! Longitudinal Birth Cohort Study** | Victoria, Australia | R | N/A | Expectant mothers and their newborns in antenatal and general practice clinics living in the Barwon-South Western (BSW) region | 2 |
| **24. Oral HPV in Indigenous South Australians cohort study** | South Australia, Australia | U, R | 2018-2019 | Aboriginal and/or Torres Strait Islanders who are older than 18 and South Australian residents, recruited through the Aboriginal Community Controlled Health Organisations (ACCHOs) | 2 |
| **25. Dental Fluorosis Among South Australian Children** | South Australia, Australia | U, R | 2003-2004 | South Australian children aged 8-14 years who attended the school dental service (SDS) | 4 |
| **26.** Tooth Eruption in Infants Cohort Study | Victoria, Australia | U | N/A | Children between 6 months and 2 years old attending three long-day childcare centres in Melbourne, three or more days per week | 1 |
| **27.** Sugar–starch combinations in food and the relationship to dental caries in low-risk adolescents | Victoria, Australia | U | 1995 | Year 7 students, aged between 12 and 13 years, attending public secondary colleges in the north-west region of metropolitan Melbourne | 1 |
| **28.** Dental caries in Taranaki Adolescents Cohort Study | Taranaki, New Zealand | N/A | 2003 | Children between 12 and 13 years of age | 1 |
| **29.** The New Zealand Very Low Birth Weight Study (VLBW) | New Zealand | U | 1986 | Live-born infants with birthweight less than 1500 g admitted for neonatal intensive care | 2 |
| **30.** Mater University of Queensland Study of Pregnancy (MUSP) | Queensland, Australia | U | 1981 | Pregnant women attending antenatal clinic of Mater Misericordiae Mothers' Hospital-University of Queensland | 1 |
| **31.** ASPREE Longitudinal Study of Older Persons (ALSOP) | Australia | U, R | 2012 | Healthy, independently living Australians aged 70 or more recruited by General Practitioners in their clinics | 2 |
| **32.** Oral Health at Five Years Study | South Australia, Australia | U, R | 2003 | Five-year-old children recruited from the electronic database of the School Dental Service in South Australia | 2 |
| **33.** Oral Health of Critically Ill Children Cohort Study | Victoria, Australia | U | 2011 | Children of all ages admitted to the Paediatric Intensive Care Unit (PICU) at the Royal Children's Hospital (RCH) | 1 |
| **34.** Malocclusion and Oral Health-Related Quality of Life Cohort Study | New Zealand | U, R | N/A | Patients between the ages of 10 and 17 years who were to undergo full maxillary and mandibular fixed orthodontic treatment (not involving headgear or other extraoral appliances) at private specialist orthodontic practices throughout New Zealand | 1 |
| **35.** University of Adelaide longitudinal study of growth and development | Northern Territory, Australia | R | 1950 - 1970 | Aboriginal people resident at Yuendumu, Central Australia | 2 |
| **36.** North Brisbane Children Cohort Study | Queensland, Australia | R | 2013 - 2015 | Children less than 5 years old attending an Aboriginal-owned and operated primary health care clinic in Caboolture, a northern suburb of Brisbane | 1 |
| **37.** The Household, Income and Labour Dynamics in Australia (HILDA) Survey | Australia | U, R | 2001 | Australian Residents | 1 |

Table 3 presents details about the recruited participants and the data collection waves. Among the 37 studies, 21 studies exclusively recruited infant cohorts as all or part of their target populations. In contrast, the “Concord Health and Ageing in Men Project (CHAMP)” and the “ASPREE Longitudinal Study of Older Persons (ALSOP)” enrolled participants from the oldest age demographic (>70 years). Twenty-two studies collected variables measuring oral health within a year of recruiting their participants. Moreover, 24 studies measured oral health across more than one wave. However, two of the included studies did not have oral health measurements but only investigated risk factors related to oral health.

**Table 3.** Design characteristics of original cohort studies with primary data collection presenting oral health data^4^.

| # | Age at recruitment (years) | Age at the first wave with oral health measurements (years) | Age at the last wave with oral health measurements (years) | Baseline sample size (highest reported) | Ethnicity in the Baseline (%) | Implemented intervention |
| --- | --- | --- | --- | --- | --- | --- |
| 1 | 13 | 13 | 30 | 3,925 | Australian Born Male Parent (70.6%), Australian Born Female Parent (74.3%) | - |
| 2 | 0 | 0 | 2 | 312 | N/A | - |
| 3 | 0 | 18 | [No follow-up measured oral health] | 686 | Aboriginal and Torres Strait Islander (100%) | - |
| 4 | Cohort 1: N/A | N/A | [No follow-up] | >600 | European Descent (~100%) | - |
|  | Cohort 2: 3-5 | 3-5 | 12-14 | 654 |  |  |
|  | Cohort 3: 0-2 | 0-2 | 13-16 | 436 |  |  |
| 5 | 0 | 7 | [No follow-up measured oral health] | 1,265 | Caucasian (93.3%), Non-Caucasian (6.7%) | - |
| 6 | 70≤ | 80≤ | 84≤ | 1,705 | Born in Australia (49.8%), Born in UK (4.6%), Born in Italy (19.6%), Born in Greece (3.81%), Other (22.17%) | - |

<sup>4</sup> In case of concurrent recruitment of parents, age and sample size of the children are reported

|  |  |  |  |  |  |  |
| --- | --- | --- | --- | --- | --- | --- |
| 7 | 0 | 5 | 45 | 1,037 | White (92.5%), Māori (7.5%) | - |
| 8 | 0 | 0 | 3 | 2,904 | Born in Australia (73.6%), Other (26.4%) | - |
| 9 | 0 | 1.5 | 9 | 159 | Aboriginal (100%) | - |
| 10 | Birth cohort (0-1) | 2-3 | 16-17 | 5,107 | Born in Australia (98.6%), Other (1.4%)<br>/<br>Indigenous (4.1%), Non-Indigenous (95.9%) | - |
|  | Kindergarten cohort (4-5) | 4-5 | 20-21 | 4,983 |  |  |
| 11 | Birth cohort (0.5-2) | 0.5-2 | 13.5-15 | 1,687 | Aboriginal and Torres Strait Islander (100%) | - |
|  | Kindergarten cohort (3.5-5) | 3.5-5 | 16.5-18 |  |  |  |
| 12 | 0 | 4 | 6 | 1,398 | Maternal: Samoan (47.2%), Cook Island (16.9%), Tongan (21%), Other Pacific (7.7%), non-Pacific [With Pacific father] (7.2%) | - |
| 13 | 0 | 0.5 | 7 | 1,052 | Mother born in Australia (74%),<br>Mother not Born in Australia (26%) | Application of Casein phosphopeptide - amorphous calcium phosphate (CPP/ACP) paste and chlorhexidine gel, Teeth brushing instructions |
| 14 | SADLS I: ≥60 | ≥60 | ≥71 | 1,650 | N/A | - |
|  | SADLS II: ≥60 | ≥60 | ≥62 | 810 |  |  |
| 15 | 0 | 0 | 5 | 2,181 | Mother born in Australia, New Zealand, or UK (73%),<br>Mother born in other Asian countries (11.4%),<br>Mother born in India (8.9%), Other (6.7%)<br>/<br>Indigenous (2.5%),<br>Non-Indigenous (97.5%) | - |
| 16 | ≥45 | ≥45 | [No follow-up measured oral health] | 267,357 | Aboriginal and Torres Strait Islander (0.7%)<br>/<br>Born in Australia (74.9%) | - |
| 17 | CFS Mark 1/A: 5-15 | 5-15 | 8-18 | 9,980 | N/A | - |
|  | CFS Mark 1/B: 5-12 | 5-12 | 8-15 | 10,695 |  |  |
|  | CFS Mark 2: 5-17 | 5-17 | [No follow-up] | 6,259 |  |  |
| 18 | 0 (18–20 weeks of gestation) | 6 | [No follow-up measured oral health] | 500 | N/A | - |
| 19 | 0 | 1 Month | 5 | 467 | Mother born in Australia (65.3%) | - |
| 20 | 0 | 4-7 | [No follow-up measured oral health] | 6,853 | European (62.3%), Māori (18.3%), Pacific Peoples (17%), Asian (15.9%), Middle Eastern/Latin American/African (2.4%), Other (0.6%) / Born in New Zealand (64.1%), Born in Australia (1.8%), Born in Other Oceania (10.5%), Born in Asia (11.6%), Born in Europe (6.7%), Born in Africa (2.7%), Born in The Americas (1.7%), Born in Middle East (0.8%) | - |
| 21 | 0 | 3-4 | [No follow-up measured oral health] | 1,036 | Of 935 participants: Mother born in Australia (38.4%), Mother born in China (4.5%), Mother born in Vietnam (12.3%), Mother born in Other Asia (11.2%), Mother born in Middle East/Africa (10.3%), Others (12.8%) | - |
| 22 | 0 | 2 | 7 | 448 | Aboriginal (100%) | Dental care to pregnant mothers, Fluoride varnish applied at child ages 6, 12 and 18 months, Anticipatory guidance, Motivational interviewing |
| 23 | 0 | [No Oral Health Measurement, Only Risk Factors] | [No Oral Health Measurement, Only Risk Factors] | 458 | N/A | - |
| 24 | 18+ | 18+ | 20+ | 1,011 | Aboriginal and/or<br>Torres Strait Islanders<br>(100%) | - |
| 25 | 8 - 13 | 8 - 13 | 16 - 21 | 677 | N/A | - |
| 26 | 0.5 - 2 | 0.5 - 2 | 1 – 2.5 | 21 | N/A | - |
| 27 | 12 - 13 | 12 - 13 | 14 - 15 | 645 | Anglo-Saxon (~50%) | - |
| 28 | 13 | 13 | 16 | 430 | Māori (20.5%), Non-Māori (79.5%) | - |
| 29 | 0 | 26 - 30 | [No follow-up<br>measured oral<br>health] | 413 | N/A | - |
| 30 | 0 | 5 | [No follow-up<br>measured oral<br>health] | 7,351 | N/A | - |
| 31 | 70+ | 71+ | 75+ | 14,892 | Born outside Australia<br>(24.4%) | - |
| 32 | 5 | 5 | [No follow-up] | 1,398 | Indigenous (2%), Non-Indigenous (98%) | - |
| 33 | 0 - 14 | 0 - 14 | (0 – 14) + a few days | 46 | N/A | - |
| 34 | 10 - 17 | 10 - 17 | 14 - 21 | 174 | European (81.6%),<br>Non-European (18.4%) | - |
| 35 | Any Age | Any Age | [No follow-up<br>measured oral<br>health] | 446 | Aboriginal (100%) | - |
| 36 | 0 - 5 | 0 - 5 | [No follow-up] | 180 | Indigenous (100%) | - |
| 37 | Any Age | [No Oral Health<br>Measurement, Only<br>Risk Factors] | [No Oral Health<br>Measurement, Only<br>Risk Factors] | 13,969 | N/A | - |

Among the 26 studies that reported on ethnicity in their baseline samples, 15 identified and reported their Indigenous populations, including Aboriginal and Torres Strait Islanders in Australia, as well as Māori and Pacific Islanders in New Zealand. Only seven studies in Australia and one study in New Zealand exclusively recruited these populations. No cohort study was identified that recruited exclusively from Culturally and Linguistically Diverse (CALD) communities.

Figures 4 and 5 present the baseline years and sample sizes of the identified primary cohort studies, respectively. A total of 25 cohort studies began between 2000 and 2015, indicating a peak period. The median baseline sample size across these studies is 1,036 participants, with the 45 and Up study being the largest, enrolling 267,357 participants across New South Wales, Australia.

While cohort studies are, by definition, observational in nature, two studies that involved interventions with participants were included: the Queensland Birth Cohort Study and the South Australian Aboriginal Birth Cohort Study (SAABC). The Queensland birth cohort study involved applying Casein phosphopeptide-amorphous calcium phosphate (CPP/ACP) paste, chlorhexidine gel, and teeth brushing instructions to a subgroup of participants through home visits and telephone calls (47). The SAABC also included dental care for pregnant mothers, fluoride varnish applied at ages 6, 12, and 18 months, anticipatory guidance, and motivational interviewing (48).

### 3.2 Independent Data Linkage Studies

Table 4 presents details of independent data linkage studies reporting oral health-related data. All these studies utilised hospital records as part of their analysed data. Seven studies were conducted in Australia and three in New Zealand. Five studies drew data from urban settings, while another five incorporated data from both urban and rural settings. Five of these studies had sample sizes larger than 20,000, with the highest being 68,543 participants (49). Only one study exclusively targeted Indigenous populations (the Māori and Pacific children in New Zealand) (50).

**Table 4.**
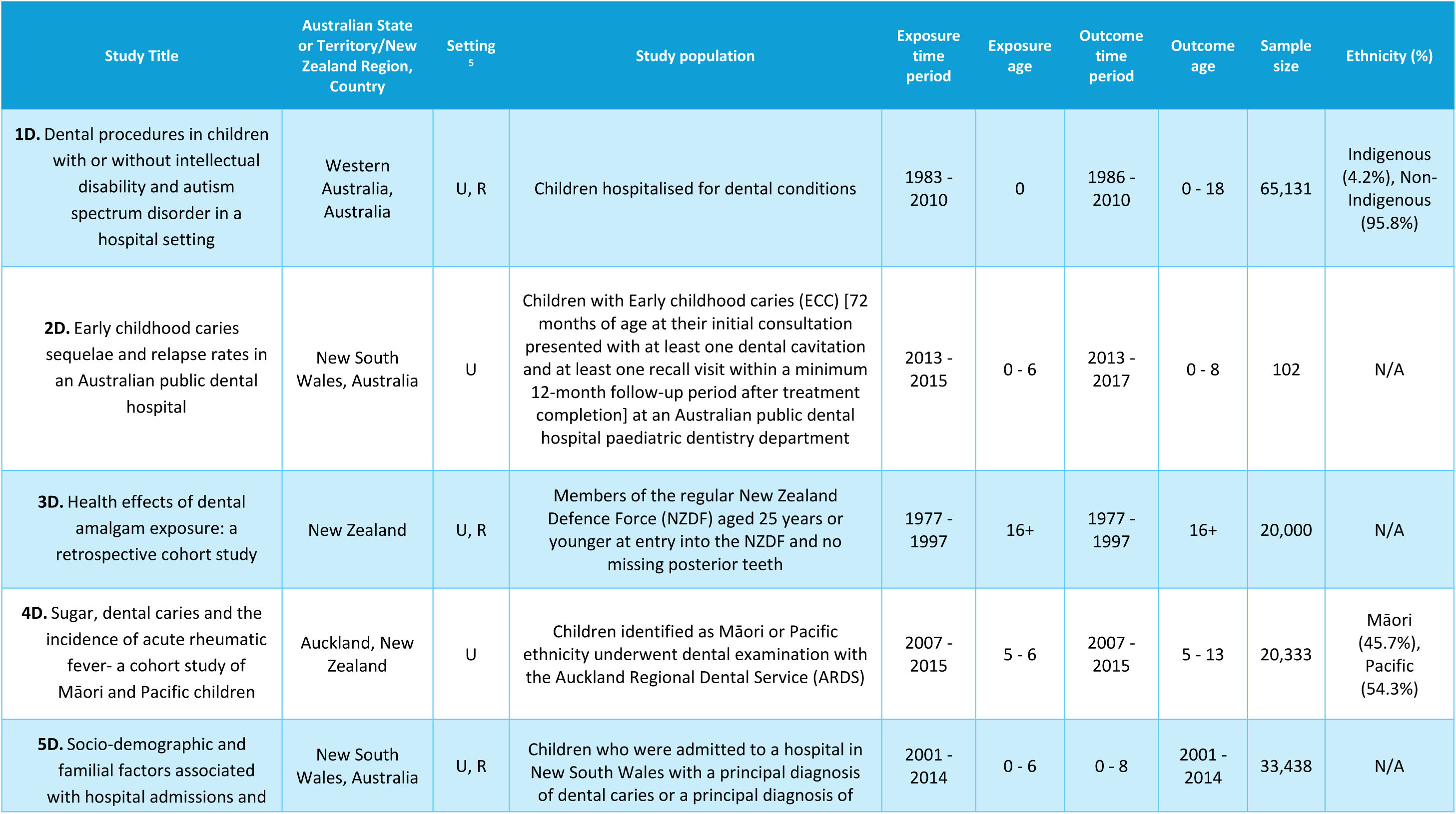
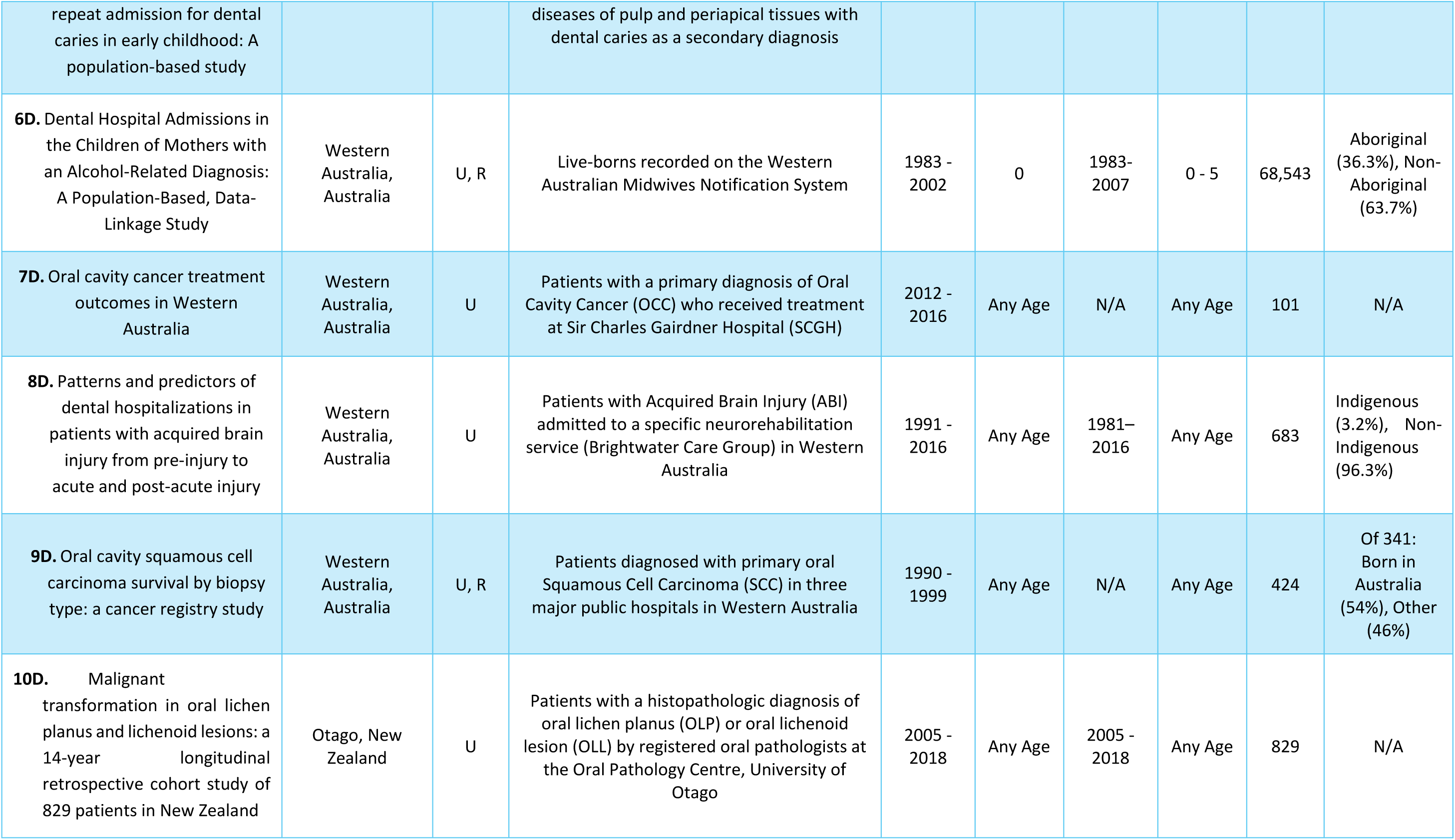
Characteristics of independent data linkage studies presenting oral health data.

| Study Title | Australian State or Territory/New Zealand Region, Country | Setting<sup>5</sup> | Study population | Exposure time period | Exposure age | Outcome time period | Outcome age | Sample size | Ethnicity (%) |
| --- | --- | --- | --- | --- | --- | --- | --- | --- | --- |
| **1D.** Dental procedures in children with or without intellectual disability and autism spectrum disorder in a hospital setting | Western Australia, Australia | U, R | Children hospitalised for dental conditions | 1983 - 2010 | 0 | 1986 - 2010 | 0 - 18 | 65,131 | Indigenous (4.2%), Non-Indigenous (95.8%) |
| **2D.** Early childhood caries sequelae and relapse rates in an Australian public dental hospital | New South Wales, Australia | U | Children with Early childhood caries (ECC) [72 months of age at their initial consultation presented with at least one dental cavitation and at least one recall visit within a minimum 12-month follow-up period after treatment completion] at an Australian public dental hospital paediatric dentistry department | 2013 - 2015 | 0 - 6 | 2013 - 2017 | 0 - 8 | 102 | N/A |
| **3D.** Health effects of dental amalgam exposure: a retrospective cohort study | New Zealand | U, R | Members of the regular New Zealand Defence Force (NZDF) aged 25 years or younger at entry into the NZDF and no missing posterior teeth | 1977 - 1997 | 16+ | 1977 - 1997 | 16+ | 20,000 | N/A |
| **4D.** Sugar, dental caries and the incidence of acute rheumatic fever- a cohort study of Māori and Pacific children | Auckland, New Zealand | U | Children identified as Māori or Pacific ethnicity underwent dental examination with the Auckland Regional Dental Service (ARDS) | 2007 - 2015 | 5 - 6 | 2007 - 2015 | 5 - 13 | 20,333 | Māori (45.7%), Pacific (54.3%) |
| **5D.** Socio-demographic and familial factors associated with hospital admissions and | New South Wales, Australia | U, R | Children who were admitted to a hospital in New South Wales with a principal diagnosis of dental caries or a principal diagnosis of | 2001 - 2014 | 0 - 6 | 0 - 8 | 2001 - 2014 | 33,438 | N/A |
| repeat admission for dental caries in early childhood: A population-based study |  |  | diseases of pulp and periapical tissues with dental caries as a secondary diagnosis |  |  |  |  |  |  |
| **6D.** Dental Hospital Admissions in the Children of Mothers with an Alcohol-Related Diagnosis: A Population-Based, Data-Linkage Study | Western Australia, Australia | U, R | Live-borns recorded on the Western Australian Midwives Notification System | 1983 - 2002 | 0 | 1983-2007 | 0 - 5 | 68,543 | Aboriginal (36.3%), Non-Aboriginal (63.7%) |
| **7D.** Oral cavity cancer treatment outcomes in Western Australia | Western Australia, Australia | U | Patients with a primary diagnosis of Oral Cavity Cancer (OCC) who received treatment at Sir Charles Gairdner Hospital (SCGH) | 2012 - 2016 | Any Age | N/A | Any Age | 101 | N/A |
| **8D.** Patterns and predictors of dental hospitalizations in patients with acquired brain injury from pre-injury to acute and post-acute injury | Western Australia, Australia | U | Patients with Acquired Brain Injury (ABI) admitted to a specific neurorehabilitation service (Brightwater Care Group) in Western Australia | 1991 - 2016 | Any Age | 1981–2016 | Any Age | 683 | Indigenous (3.2%), Non-Indigenous (96.3%) |
| **9D.** Oral cavity squamous cell carcinoma survival by biopsy type: a cancer registry study | Western Australia, Australia | U, R | Patients diagnosed with primary oral Squamous Cell Carcinoma (SCC) in three major public hospitals in Western Australia | 1990 - 1999 | Any Age | N/A | Any Age | 424 | Of 341: Born in Australia (54%), Other (46%) |
| **10D.** Malignant transformation in oral lichen planus and lichenoid lesions: a 14-year longitudinal retrospective cohort study of 829 patients in New Zealand | Otago, New Zealand | U | Patients with a histopathologic diagnosis of oral lichen planus (OLP) or oral lichenoid lesion (OLL) by registered oral pathologists at the Oral Pathology Centre, University of Otago | 2005 - 2018 | Any Age | 2005 - 2018 | Any Age | 829 | N/A |

### 3.3 Oral Health Measurements

Oral health measurements derived from the identified cohort studies are illustrated as a bipartite network in Figure 6. Ignoring the variations among similar variables across studies, we identified a total of 70 distinct oral health measurements, categorising them into self-reported (n = 22) and clinical (n = 48) variables. Regarding clinical data, the DMFT (Decayed, Missing, Filled Teeth)/dmft and DMFS (Decayed, Missing, Filled Surfaces)/dmfs indices emerged as the most commonly utilised measurements, employed in 20 studies, followed by “Dental Caries Presence (Cavitation/White Spot Lesion)” (n = 6) and “Oral Microbiota” (n = 6). In terms of self-reported oral health measurements, “Self-rated Oral Health” (n = 11), “Dento-facial Pain” (n = 6), and “Teeth Number/Missing Tooth/Tooth Loss” (n = 5) were the most frequently reported. The DMHDS recorded the highest number of oral health measurements (n = 16), followed by CHAMP (n = 13) and the South Australian Dental Longitudinal Study (SADLS) (n = 12).

### 3.4 Explanatory Factors and Outcomes

Figure 7 presents the EGM of the explanatory factors and outcomes linked together within publications of the identified oral health-related cohort studies. A comprehensive total of 38 themes pertaining to explanatory factors and 32 themes related to outcomes have been identified. The most extensively studied connections were “Sociodemographic/Economic/Cultural Factors (Including Access and Use of Dental Services)” to “Dental/Oral Condition: Dental Caries/Tooth Loss” (n = 25), “Oral Hygiene, Behaviours and Beliefs” to “Dental/Oral Condition: Dental Caries/Tooth Loss” (n = 22), “Diet and Nutrient Intake” to “Dental/Oral Condition: Dental Caries/Tooth Loss” (n = 16), and “Parental Health and Habits” to “Dental/Oral Condition: Dental Caries/Tooth Loss” (n = 12).

Figures 8 and 9 present the examined explanatory factors and outcomes separately. Regarding explanatory factors, “Sociodemographic/Economic/Cultural Factors (Including Access and Use of Dental Services)” (n = 41), “Oral Hygiene, Behaviours and Beliefs” (n = 25), “Diet and Nutrient Intake” (n = 17), “Parental Health and Habits” (n = 16), and “Medical Conditions/Treatments/Presentations: Other” (n = 16) emerged as the most studied. In terms of outcomes, “Dental/Oral Condition: Dental Caries/Tooth Loss” (n = 28), “Medical Conditions/Treatments/Presentations: Other” (n = 10), “Dental/Oral Condition: Developmental Dental Defects” (n = 7), “Perception of Personal Oral Health and Needs (Including Oral Health Quality of Life)” (n = 7), and “Oral Hygiene, Behaviours and Beliefs” (n = 6) were the most investigated.

## 4. Discussion

This review aims to provide an overview of cohort studies on oral health throughout the lifespan in Australia and New Zealand, which is essential for coordinated research and policy development to address oral health challenges in the region. A total of 37 cohort studies with primary data collection and 10 independent data linkage studies were identified, resulting in 226 publications. The geographical distribution, general features, and design aspects of the identified studies were summarised. Seventy oral health measurements used in these studies were identified and categorised. Moreover, based on the publications from the identified cohort studies, an EGM was displayed to depict the examined connections between 38 themes for explanatory factors and 32 themes for outcomes in these studies.

### 4.1 Primary Cohort Studies

This review highlights a clear upward trend in the identified cohort studies conducted in the region and the number of resultant publications over recent years and decades. This trend can probably be attributed to increased public awareness and the implementation of national oral health promotion policies in both countries. Most notably, implementation of the first Australian “National Oral Health Plan” in 2004, alongside New Zealand’s “Good Oral Health for All for Life” national strategy, highlights a clear prioritisation of oral health initiatives in these nations (51, 52). However, past novel projects in the region, such as the DMHDS in New Zealand initiated in 1975, have also provided valuable insights into oral health life course trajectories, encouraging and laying the foundation for more recent research endeavours (53). Finally, development of data infrastructure facilitated access to a variety of patient records, providing an opportunity to conduct data linkage studies (54, 55).

A varied geographic distribution of the cohort studies was observed across the region, with more being identified in Australia compared to New Zealand, alongside a higher concentration of primary cohort studies in the eastern states of Australia. This disparity may simply reflect the larger populations in those areas (56, 57). However, it is important to recognise the influence of well-established institutions, such as the University of Adelaide Dental School and its affiliated oral health-focused research centre – Australian Research Centre for Population Oral Health (ARCPOH) (58). They have significantly contributed to South Australia having the highest number of primary oral health-related cohort studies among other regions, including hosting the oldest identified study, the University of Adelaide Longitudinal Study of Growth and Development (59). Nevertheless, it is essential to understand that the quantity of studies conducted does not inherently reflect their significance or value. Factors such as sample size, follow-up duration, attrition rate, and the rigour of data collection and analysis are critical in determining the value and relevance of research within the domain of life course oral health (60).

This review also identified an uneven focus across different age groups. Most of the identified studies recruited participants from childhood, primarily from birth. This focus, however, is by no means unjustified, as childhood studies facilitate longitudinal tracking into adulthood, allowing for the investigation of developmental trajectories and the impacts of early life exposures (61). Moreover, childhood is frequently targeted because early intervention yields cost-effective preventive strategies, potentially mitigating future oral health burdens (62). However, recruiting participants from their childhoods could limit the study’s reach regarding conditions that typically occur in older ages, such as periodontal disease, edentulism, and oral cancer (19, 63, 64), as it requires long-term follow-ups, which might not be feasible due to limited resources and inevitably high attrition rates (65, 66). Another emphasis in the reviewed studies was on recruiting older adults (aged over 60) (67–69). This newly emerged focus on older adults is mainly due to the increasing ageing population in Australia and New Zealand, along with concerns regarding the rising prevalence of chronic oral diseases that impact quality of life (70, 71). Although a few studies, like the DMHDS or the Longitudinal Study of Australian Children (LSAC), managed to follow their participants into their early adulthood (i.e., 18 – 25 years) (34, 72), there remains an extremely limited number of studies exclusively recruiting adult participants. The limited interest in recruiting individuals aged 18 – 45 years may be attributed to logistical difficulties linked to recruitment and retention. These challenges might primarily stem from transient lifestyles, especially in early adulthood, as well as high mobility resulting from employment changes, family dynamics, and economic factors (73, 74). The observed age-related representation gap may influence the generalisability and applicability of findings in oral health cohort studies, potentially obscuring vital risk and protective factors that emerge distinctly during these periods.

Another key point to consider is the ethnic diversity and focus of the identified cohort studies. Few identified studies investigated exclusive Indigenous populations in Australia (n = 7) and New Zealand (n = 2), with no study focusing on CALD populations, showing a clear gap and under-representation of these minority groups. This observation is crucial, particularly given the established oral health disparities affecting the Aboriginal and Torres Strait Islander populations in Australia (75), as well as the Māori and Pacific Islanders in New Zealand (76, 77). Concerning CALD communities in the region, while studies comparing their oral health status to the general population are limited, disparities in their dental care utilisation have been observed (78, 79). However, obstacles to establishing exclusive cohort studies of these minority groups, including historical mistrust and past unethical practices involving Indigenous populations, community engagement difficulties, lack of cultural safety and competency, and language and communication barriers, should be accounted for (80–83). Fortunately, Australia and New Zealand have turned their focus on the health disparities affecting minority groups, especially Indigenous populations, as shown by national and regional action plans (84–86); however, significant gaps still remain that need to be addressed.

Most identified cohort studies primarily focused on general health, with oral health being a secondary concern. Only a few studies, such as HSHK and SMILE, were originally designed to address oral health issues, as their initial protocols indicate (87, 88). While this broader focus on overall health facilitates drawing connections between systemic and oral health, it limits the depth and precision of collected oral health data, potentially leaving gaps in important outcomes and risk factors. Consequently, these studies may not fully capture the complexity of oral health issues within the studied populations.

### 4.2 Data Linkage Studies

This review identified and characterised oral health studies that rely solely on data linkage (n = 10). Half of these independent data linkage studies utilised data from Western Australia. The rise in data linkage studies in this area can be attributed to the establishment of the “WA Data Linkage System”, which has significantly facilitated the process of conducting such research (89). However, it is worth mentioning that some cohort studies with primary data collection, such as LSAC (90), have incorporated data linkage alongside other data collection efforts; however, they have not been counted as independent data linkage studies in this review.

Despite being limited in number, their distinct contributions to the field cannot be overlooked. The utilisation of hospital records and cancer registries in these identified studies enabled the investigation of rare oral and medical conditions, such as oral cancer and other epithelial disorders. The capacity to incorporate a large sample size in these investigations allowed for the examination of these outcomes, even though they occur infrequently, making them nearly impossible to be investigated through primary cohort studies (91). This increased sample size would also lead to enhanced representativeness, improving statistical power, reducing selection bias, and enhancing the generalisability of findings (92). However, the inherent limitations of data linkage studies, including potential inaccuracies in administrative datasets, inconsistencies in record linkage quality, and ethical concerns surrounding data privacy, should be acknowledged (93). Regarding the latter concern, Australia and New Zealand have developed sophisticated ethical review processes, like the Five Safes framework and the Privacy Impact Assessment (PIA), to safeguard data privacy (94, 95).

### 4.3 Oral Health Measurements

This review identified 70 oral health measurements, categorised into 48 clinically measured and 22 self-reported. However, there are many variations and overlaps among these measurements. For example, dental caries measurements included DMFS (Decayed, Missing, Filled Surfaces) (96), ICDAS (International Caries Detection and Assessment System) (97), self-reported caries experiences (98), and various non-specific measures of cavitation and white spots (99). Likewise, Oral Health-related Quality of Life (OHRQoL) was assessed through several standardised questionnaires or general inquiries, such as the Oral Health Impact Profile (OHIP-14) Scores (100), Child and Parental Perception Questionnaires (CPQ and PPQ) (101), and other self-reported questions regarding pain, function, and overall oral health. These discrepancies limit the direct comparisons and meta-analyses of study outcomes, making it challenging or impossible to synthesise more statistically robust results (102). Furthermore, employing multiple similar but non-identical tools for the same construct introduces a risk of misclassification bias (103). For instance, self-reported dental caries, when compared with clinically measured DMFT (Decayed, Missing, Filled Teeth), has been shown to underestimate the true prevalence of this condition, potentially leading to misleading conclusions about disease burden (104).

The distinction between clinical and self-reported oral health measurements illustrated in this review should also be acknowledged. Clinical data obtained from examinations and various clinical tools primarily target specific conditions such as dental caries, periodontal diseases, and other oral health issues. In contrast, self-reported data emphasise subjective experiences, including quality of life, aesthetics, pain, and other symptoms, rather than reflecting specific conditions. While clinical measurements are the traditional method for evaluating oral health status in research and offer more reliable and objective results, they demand considerable resources and can be expensive (105). Furthermore, relying solely on clinical measurements often overlooks patient-centred outcomes, thereby failing to capture a complete picture of oral health within communities (106). Therefore, integrating clinical and self-reported data in oral health cohort studies is vital for a thorough assessment of oral health, promoting a more holistic perspective on health (106, 107).

Additionally, it is important to acknowledge the development of new oral health measurements and their implementation in the identified cohort studies. Salivary biomarkers (such as immunoglobulin assays) (108), oral microbiota (including Streptococcus mutans and Lactobacilli) (109, 110), salivary pH (110), and oral flora DNA testing for Human Papillomavirus (HPV) (111) are some of the innovative measures employed by cohort studies. These investigations enable earlier detection of disease, better risk stratification, and a clearer understanding of the interaction between oral and systemic health (112, 113). However, their utility is constrained by cost, standardisation, ethical issues, participant compliance, analytical complexity, and integration challenges (113, 114).

### 4.4 Evidence and Gap Map

This review presents an evidence and gap map of the outcomes and explanatory factors examined across the identified oral health-related cohort studies, highlighting areas with varying concentrations of evidence. Dental caries is by far the most studied outcome, reflecting its status as the most common oral health condition (115). However, developmental dental defects, the next most examined oral condition, do not correlate with their prevalence. Conversely, periodontal diseases, despite being the second most prevalent oral health issue globally (116), were studied infrequently. This observation is likely due to the natural age of onset of these conditions—developmental dental defects manifesting in early life (117) and periodontal disease appearing later in life (116)—and the previously discussed age distribution of participants in the cohort studies. Regarding explanatory factors, social determinants of health were investigated most extensively, likely due to their established foundational role in shaping oral and general health outcomes and driving disparities across populations (31). Following this, oral hygiene, behaviours and beliefs, diet and nutrient intake and substance use (including smoking and alcohol consumption) were extensively employed to explain oral health outcomes, representing core behavioural and lifestyle determinants of oral health, influencing the initiation and progression of dental caries, periodontal disease, tooth loss, oral cancer, and many other oral health problems (118–120). The common characteristic of these explanatory variables is their modifiability at both individual and population levels, making them vital for developing prevention strategies and health promotion policies (121), thereby justifying their frequent use in oral health research. Finally, parental health and habits were consistently examined throughout the identified cohort studies to explain children’s oral health outcomes. Parents play a direct role in shaping their children’s oral hygiene habits, eating behaviours, and the use of dental services by modelling behaviours and creating household routines (122, 123). Moreover, parental systemic and oral health can directly impact children’s oral health through the biological transmission of cariogenic bacteria (124), the inheritance of faulty genes (125), and various other mechanisms, making them a focal point for the identified cohort studies.

Less studied areas are another highlight of this review. Certain important factors such as temporomandibular joint disorders (TMD), oral microbiota, salivary biomarkers, and cognitive and mental health appear relatively sparse on the evidence gap map. Nevertheless, they are regarded as promising areas for innovative research and clinical insights into their role within the overall context of oral health in the existing literature (126–128). Exploring these underrepresented domains may enable earlier disease detection, enhance personalised approaches to oral healthcare, and incorporate oral health more thoroughly within broader medical and public health frameworks (129, 130).

### 4.5 Strengths

To the best of our knowledge, this systematic mapping review represents the first comprehensive assessment of oral health-related cohort studies in Australia and New Zealand, incorporating all types of cohort studies across the lifespan. The strategic application of various visual and descriptive synthesis tools strengthens this systematic mapping review by enhancing data presentation, accessibility, and impact. In particular, the interactive bipartite network and evidence and gap map are innovative tools that provide a clear visualisation of the connections between cohort studies and oral health measurements, as well as the explanatory factors and outcomes, respectively, delivering statistical data without sacrificing study-level details.

### 4.6 Limitations

Despite having a comprehensive electronic database and conducting backward citation chasing, this review may have overlooked some relevant studies, particularly due to the broad eligibility criteria. The heterogeneity among included publications prevented a formal assessment of methodological quality or risk of bias. Presenting the vast number of variables collected from the identified studies required a degree of simplification and categorisation, increasing the likelihood of not fully capturing the nuances or specific definitions utilised in each study. Lastly, the inclusion of only published literature may introduce publication bias, and ongoing or unpublished cohort studies with oral health data might be missed.

### 4.7 Implications for Future Research, Policy, and Practice

This review offers a valuable guide for researchers to identify the over- and under-represented topics and populations in oral health research and to direct their future research efforts. Heavily investigated links in the EGM would present opportunities for the conduct of future systematic reviews, potentially including meta-analyses. Additionally, there is potential for conducting secondary research based on the data sharing of the identified cohort studies. This would be facilitated by establishing a regional oral health cohort registry or open-access datasets, which might enable individual-level pooled analyses (131). Given the heterogeneity between assessment and outcome measures across studies, this review underscores the necessity to prioritise the development and adoption of core outcome sets (132) and standardised methodologies for common oral conditions and key variables in future research. Furthermore, the identified under-represented age groups (i.e. middle-aged adults), ethnicities (i.e. Indigenous and migrant communities), regions, oral conditions, and explanatory factors should be the focus of future research. Finally, further development and ethical use of data linkage methodologies, along with efforts to enhance the quality and interoperability of administrative datasets, should be promoted.

The comprehensive overview of current oral health cohort studies can effectively inform policymakers on resource allocation and strategic investments in areas with the greatest research needs or potential impact. A key area for such investment is the ongoing and enhanced support for cohort studies focused on ethnic minorities who face oral health disparities. This review also underscores the substantial existing evidence that should further inform evidence-based decision-making regarding population-level oral health policy development. Lastly, identified cohort studies are essential for supplying the longitudinal data needed to comprehend disease trends, assess the success of oral health promotion initiatives, and track advancements toward achieving national oral health objectives.

This review presents notable implications for clinical practice as well. It promotes greater clinical awareness of oral health issues, particularly in under-explored areas. Identification of key risk factors for oral diseases will enable earlier and more targeted preventive interventions, particularly in childhood and adolescence. This review also highlights the intricate relationship between oral and systemic health, underscoring the necessity of integrated care approaches to oral health. The findings highlight ongoing disparities in oral health outcomes among Indigenous and socioeconomically disadvantaged populations, underscoring the need for culturally sensitive and equitable care models. Overall, the evidence supports the development of more personalised, inclusive, and effective oral healthcare strategies.

## 5. Conclusion

This systematic mapping review highlights the existence of a substantial number of cohort studies related to oral health in Australia and New Zealand, with significant variations in their key characteristics, participant demographics, oral health measurements, and the investigated links of explanatory factors to outcomes. While over-represented study populations, oral health measurements, and determinant-outcome associations present opportunities for further secondary research, such as individual-level pooled analysis, under-represented areas should become the focus of primary research moving forward. Therefore, this review serves as a roadmap for researchers, as well as policymakers and clinicians, in making evidence-based decisions to improve oral health in the region.

## Data Availability

All data produced in the present study are available upon reasonable request to the authors

## Acknowledgments

We acknowledge the assistance of Ms. Kanchana Ekanayake, the School of Health Sciences librarian at Western Sydney University, in creating and refining the search strategy. Moreover, the role of Ms. Navodya Selvaratnam and Ms. Mariam Al Asaad in selection and data extraction process is appreciated.

---

**Figure 2.** Publication trends of 226 articles included from oral health-related cohort studies

https://parsa199978.github.io/Figure-2/

**Figure 4.** Baseline year of oral health-related cohort studies with primary data collection (n = 37)

https://parsa199978.github.io/Figure-4/

**Figure 5.** Baseline sample size of oral health-related cohort studies with primary data collection (n = 37)

https://parsa199978.github.io/Figure-5/

**Figure 6.** Interactive bipartite network of oral health measurements collected within the oral health-related cohort studies with primary data collection (n = 37) and independent data linkage studies (n = 10)

https://parsa199978.github.io/Figure-6/

**Figure 7.** Interactive Evidence Gap Map (EGM) of investigated outcomes and explanatory factors in the oral health-related cohort studies with primary data collection (n = 37) and independent data linkage studies (n = 10)

https://parsa199978.github.io/Figure-7/

**Figure 8.** Interactive pie chart of investigated explanatory factors in the oral health-related cohort studies with primary data collection (n = 37) and independent data linkage studies (n = 10)

https://parsa199978.github.io/Figure-8

**Figure 9.** Interactive pie chart of investigated outcomes in the oral health-related cohort studies with primary data collection (n = 37) and independent data linkage studies (n = 10)

https://parsa199978.github.io/Figure-9/

## 6. Appendices

**Appendix 1** https://github.com/Parsa199978/Appendix-1/raw/refs/heads/main/Appendix%201.docx

**Appendix 2** https://github.com/Parsa199978/Appendix-2/raw/refs/heads/main/Appendix%202.docx

**Appendix 3** https://github.com/Parsa199978/Appendix-3/raw/refs/heads/main/Appendix%203.docx

**Appendix 4** https://github.com/Parsa199978/Appendix-4/raw/refs/heads/main/Appendix%204.xlsx

**Appendix 5** https://github.com/Parsa199978/Appendix-5/raw/refs/heads/main/Appendix%205.docx

## Footnotes

1 Data linkage studies are marked with asterisks (*)

2 Urban: U, Rural: R

3iii Number of published articles presenting oral health data. Please note that articles presenting data from more than one cohort study have been counted towards the total number of published articles for all of the respective cohort studies.

4 In case of concurrent recruitment of parents, age and sample size of the children are reported

5 Urban: U, Rural: R

## Notes

### Competing Interest Statement

The authors have declared no competing interest.

### Clinical Protocols

https://osf.io/2948n

### Funding Statement

This study did not receive any funding

