## Appendix 2 for "Oral Health Research Across the Lifespan: A Systematic Mapping Review of Cohort Studies in Australia and New Zealand"

**First search 9/12/23**

Ovid MEDLINE(R) ALL <1946 to December 07, 2023>

1	Oral Health/ or exp Mouth Diseases/ or exp Dental Care/ or exp Mouth Neoplasms/ or exp Dental Caries/ or exp Tooth Injuries/ or exp Periodontitis/ or exp Dentition, Mixed/ or exp Dentition/ or exp Dentition, Permanent/ or exp Xerostomia/ or exp Malocclusion, Angle Class II/ or exp Malocclusion/ or exp Malocclusion, Angle Class I/ or exp Malocclusion, Angle Class III/ or exp Toothache/ or exp DMF Index/ or exp Tooth Diseases/	607854

2	(oral health* or dental health* or oral disease* or dental disease* or dental caries* or tooth decay* or oral trauma* or gum disease* or periodont* or gingiv* or oral hygiene* or mouth diseas* or dental car* or mouth tumo?r or Mouth Neoplasm* or mouth cancer* or Oral cancer* or Oral traum* or dental visit* or tooth loss* or edent* or dental status* or tooth* or dental pain* or dentition* or dental* or malocclusion* or teeth* or xerostomia* or tooth diseas* or dental care* or Tooth Injur* or toothache*).mp.	802132

3	1 or 2	967100

4	exp Cohort Studies/	2548195

5	Observational Study/	149535

6	(Cohort* or follow up* or follow-up* or longitudinal* or prospective* or retrospective* or observational*).mp.	4213769

7	4 or 5 or 6	4213769

8	exp Australia/ or new Zealand/	210069

9	(Australia* or oceani* or "New South Wales" or Sydney or Queensland or Brisbane or Victoria or Melbourne or "Australian Capital Territory" or Canberra or "Northern Territory" or Darwin or "South Australia" or Adelaide or Tasmania or Hobart or "Western Australia" or Perth or "New Zealand" or Christchurch or Dunedin or wellington* or Auckland).mp.	338002

10	8 or 9	338002

11	3 and 7 and 10	**1512**

**Embase Classic <1947 to 1973> Part 1 of 2**

**Embase <1974 to 2023 December 13>**

1	mouth disease/ or mouth injury/ or mouth lesion/ or mouth tumor/ or mouth ulcer/ or tooth disease/ or exp mouth tumor/ or mouth cancer/ or dental caries/ or tooth injury/ or tooth disease/ or periodontitis/ or mouth inflammation/ or periodontal disease/ or dentition/ or View results by resource xerostomia/ or angle class III malocclusion/ or malocclusion/ or angle class II malocclusion/ or angle class I malocclusion/ or tooth pain/ or exp dmf index/	382277

2	(oral health* or dental health* or oral disease* or dental disease* or dental caries* or tooth decay* or oral trauma* or gum disease* or periodont* or gingiv* or oral hygiene* or mouth diseas* or dental car* or mouth tumo?r or Mouth Neoplasm* or mouth cancer* or Oral cancer* or Oral traum* or dental visit* or tooth loss* or edent* or dental status* or tooth* or dental pain* or dentition* or dental* or malocclusion* or teeth* or xerostomia* or tooth diseas* or dental care* or Tooth Injur* or toothache*).mp.	825887

3	1 or 2	939673

4	cohort analysis/ or observational study/ or follow up/ or longitudinal study/ or prospective study/ or retrospective study/ or observational study/ or observational method/	4619409

5	(Cohort* or follow up* or follow-up* or longitudinal* or prospective* or retrospective* or observational*).mp.	6372187

6	4 or 5	6372187

7	exp Australia/ or New Zealand/	258050

8	(Australia* or oceani* or "New South Wales" or Sydney or Queensland or Brisbane or Victoria or Melbourne or "Australian Capital Territory" or Canberra or "Northern Territory" or Darwin or "South Australia" or Adelaide or Tasmania or Hobart or "Western Australia" or Perth or "New Zealand" or Christchurch or Dunedin or wellington* or Auckland).mp.	461361

9	7 or 8	461361

10	3 and 6 and 9	1996

<https://ezproxy.library.usyd.edu.au/login?url=http://ovidsp.ovid.com/ovidweb.cgi?T=JS&NEWS=N&PAGE=main&SHAREDSEARCHID=3A45yJ4RdRTv5WZplGRg2GfDdpXCIayVKhjKBRtab1uCiZbYGD26skbLUsgbazOEj>

**Scopus**

**1798 documents**

( TITLE-ABS-KEY ( "oral health*" OR "dental health*" OR "oral disease*" OR "dental disease*" OR "dental caries*" OR "tooth decay*" OR "oral trauma*" OR "gum disease*" OR periodont* OR gingiv* OR "oral hygiene*" OR "mouth diseas*" OR "dental car*" OR "mouth tumo*r" OR "Mouth Neoplasm*" OR "mouth cancer*" OR "Oral cancer*" OR "Oral traum*" OR "dental visit*" OR "tooth loss*" OR edent* OR "dental status*" OR tooth* OR "dental pain*" OR dentition* OR dental* OR malocclusion* OR teeth* OR xerostomia* OR "tooth diseas*" OR "dental care*" OR "Tooth Injur*" OR toothache* ) AND TITLE-ABS-KEY ( cohort* OR "follow up*" OR follow-up* OR longitudinal* OR prospective* OR retrospective* OR observational* ) AND TITLE-ABS-KEY ( australia* OR oceani* OR "New South Wales" OR sydney OR queensland OR brisbane OR victoria OR melbourne OR "Australian Capital Territory" OR canberra OR "Northern Territory" OR darwin OR "South Australia" OR adelaide OR tasmania OR hobart OR "Western Australia" OR perth OR "New Zealand" OR christchurch OR dunedin OR wellington* OR auckland ) )

**Web of Science**

**1031 results**

<https://www.webofscience.com/wos/woscc/summary/2cacd2c4-0e88-4919-90e0-9ee8dd5e31b0-bd0f8128/relevance/1>

| Search Query | Results |
| --- | --- |
| "oral health*" OR "dental health*" OR "oral disease*" OR "dental disease*" OR "dental caries*" OR "tooth decay*" OR "oral trauma*" OR "gum disease*" OR periodont* OR gingiv* OR "oral hygiene*" OR "mouth diseas*" OR "dental car*" OR "mouth tumo*r" OR "Mouth Neoplasm*" OR "mouth cancer*" OR "Oral cancer*" OR "Oral traum*" OR "dental visit*" OR "tooth loss*" OR edent* OR "dental status*" OR tooth* OR "dental pain*" OR dentition* OR dental* OR malocclusion* OR teeth* OR xerostomia* OR "tooth diseas*" OR "dental care*" OR "Tooth Injur*" OR toothache* (Topic) | 540457 |
| cohort* OR "follow up*" OR follow-up* OR longitudinal* OR prospective* OR retrospective* OR observational* (Topic) | 4028804 |
| australia* OR oceani* OR "New South Wales" OR sydney OR queensland OR brisbane OR victoria OR melbourne OR "Australian Capital Territory" OR canberra OR "Northern Territory" OR darwin OR "South Australia" OR adelaide OR tasmania OR hobart OR "Western Australia" OR perth OR "New Zealand" OR christchurch OR dunedin OR wellington* OR auckland (Topic) | 763789 |
| #3 AND #2 AND #1 | **1031** |

Total -6340

Duplicate 2140

4200 without Cinahl

| **Print Search History** |  |  |
| --- | --- | --- |
|  | Wednesday, December 20, 2023 9:55:33 PM |  |
| **#** | **Query** | **Results** |
| S23 | S14 AND S18 AND S22 | 1,084 |
| S22 | S19 OR S20 OR S21 | 225,478 |
| S21 | australia* OR oceani* OR "New South Wales" OR sydney OR queensland OR brisbane OR victoria OR melbourne OR "Australian Capital Territory" OR canberra OR "Northern Territory" OR darwin OR "South Australia" OR adelaide OR tasmania OR hobart OR "Western Australia" OR perth OR "New Zealand" OR christchurch OR dunedin OR wellington* OR auckland | 225,478 |
| S20 | (MH "New Zealand") | 31,900 |
| S19 | (MH "Australia+") OR (MH "Western Australia") OR (MH "South Australia") | 129,047 |
| S18 | S15 OR S16 OR S17 | 1,594,596 |
| S17 | Cohort* or follow up* or follow-up* or longitudinal* or prospective* or retrospective* or observational* | 1,274,150 |
| S16 | (MH "Nonexperimental Studies+") OR (MH "Prospective Studies+") | 924,752 |
| S15 | (MH "Prospective Studies+") OR (MH "Concurrent Prospective Studies") OR (MH "Nonconcurrent Prospective Studies") | 531,655 |
| S14 | S1 OR S2 OR S3 OR S4 OR S5 OR S6 OR S7 OR S8 OR S9 OR S10 OR S11 OR S12 OR S13 | 219,839 |

Total with duplicate -7424

Duplicate 2745

Total with duplicate 4679

Update search strategies - 16/5/25

Ovid MEDLINE(R) ALL <1946 to May 16, 2025>

1	Oral Health/ or exp Mouth Diseases/ or exp Dental Care/ or exp Mouth Neoplasms/ or exp Dental Caries/ or exp Tooth Injuries/ or exp Periodontitis/ or exp Dentition, Mixed/ or exp Dentition/ or exp Dentition, Permanent/ or exp Xerostomia/ or exp Malocclusion, Angle Class II/ or exp Malocclusion/ or exp Malocclusion, Angle Class I/ or exp Malocclusion, Angle Class III/ or exp Toothache/ or exp DMF Index/ or exp Tooth Diseases/	633690

2	(oral health* or dental health* or oral disease* or dental disease* or dental caries* or tooth decay* or oral trauma* or gum disease* or periodont* or gingiv* or oral hygiene* or mouth diseas* or dental car* or mouth tumo?r or Mouth Neoplasm* or mouth cancer* or Oral cancer* or Oral traum* or dental visit* or tooth loss* or edent* or dental status* or tooth* or dental pain* or dentition* or dental* or malocclusion* or teeth* or xerostomia* or tooth diseas* or dental care* or Tooth Injur* or toothache*).mp.	849053

3	1 or 2	1019995

4	exp Cohort Studies/	2753883

5	Observational Study/	176624

6	(Cohort* or follow up* or follow-up* or longitudinal* or prospective* or retrospective* or observational*).mp.	4641842

7	4 or 5 or 6	4641842

8	exp Australia/ or new Zealand/	222343

9	(Australia* or oceani* or "New South Wales" or Sydney or Queensland or Brisbane or Victoria or Melbourne or "Australian Capital Territory" or Canberra or "Northern Territory" or Darwin or "South Australia" or Adelaide or Tasmania or Hobart or "Western Australia" or Perth or "New Zealand" or Christchurch or Dunedin or wellington* or Auckland).mp.	360918

10	8 or 9	360918

11	3 and 7 and 10	1616

12	limit 11 to yr="2023 -Current"	**215**

**Cinahl**

| **#** | **Query** | **Limiters/Expanders** | **Last Run Via** | **Results** |
| --- | --- | --- | --- | --- |
| S24 | S14 AND S18 AND S22 | Limiters - Publication Date: 20230101-20251231 | Interface - EBSCOhost Research Databases | 53 |
|  |  | Expanders - Apply equivalent subjects | Search Screen - Advanced Search |  |
|  |  | Search modes - Proximity | Database - CINAHL Complete |  |
| S23 | S14 AND S18 AND S22 | Expanders - Apply equivalent subjects | Interface - EBSCOhost Research Databases | 606 |
|  |  | Search modes - Proximity | Search Screen - Advanced Search |  |
|  |  |  | Database - CINAHL Complete |  |
| S22 | S19 OR S20 OR S21 | Expanders - Apply equivalent subjects | Interface - EBSCOhost Research Databases | 209,112 |
|  |  | Search modes - Proximity | Search Screen - Advanced Search |  |
|  |  |  | Database - CINAHL Complete |  |
| S21 | australia* OR oceani* OR "New South Wales" OR sydney OR queensland OR brisbane OR victoria OR melbourne OR "Australian Capital Territory" OR canberra OR "Northern Territory" OR darwin OR "South Australia" OR adelaide OR tasmania OR hobart OR "Western Australia" OR perth OR "New Zealand" OR christchurch OR dunedin OR wellington* OR auckland | Expanders - Apply equivalent subjects | Interface - EBSCOhost Research Databases | 209,112 |
|  |  | Search modes - Proximity | Search Screen - Advanced Search |  |
|  |  |  | Database - CINAHL Complete |  |
| S20 | (MH "New Zealand") | Expanders - Apply equivalent subjects | Interface - EBSCOhost Research Databases | 32,979 |
|  |  | Search modes - Proximity | Search Screen - Advanced Search |  |
|  |  |  | Database - CINAHL Complete |  |
| S19 | (MH "Australia+") OR (MH "Western Australia") OR (MH "South Australia") | Expanders - Apply equivalent subjects | Interface - EBSCOhost Research Databases | 135,687 |
|  |  | Search modes - Proximity | Search Screen - Advanced Search |  |
|  |  |  | Database - CINAHL Complete |  |
| S18 | S15 OR S16 OR S17 | Expanders - Apply equivalent subjects | Interface - EBSCOhost Research Databases | 974,490 |
|  |  | Search modes - Proximity | Search Screen - Advanced Search |  |
|  |  |  | Database - CINAHL Complete |  |
| S17 | Cohort* or follow up* or follow-up* or longitudinal* or prospective* or retrospective* or observational* | Expanders - Apply equivalent subjects | Interface - EBSCOhost Research Databases | 2 |
|  |  | Search modes - Proximity | Search Screen - Advanced Search |  |
|  |  |  | Database - CINAHL Complete |  |
| S16 | (MH "Nonexperimental Studies+") OR (MH "Prospective Studies+") | Expanders - Apply equivalent subjects | Interface - EBSCOhost Research Databases | 974,490 |
|  |  | Search modes - Proximity | Search Screen - Advanced Search |  |
|  |  |  | Database - CINAHL Complete |  |
| S15 | (MH "Prospective Studies+") OR (MH "Concurrent Prospective Studies") OR (MH "Nonconcurrent Prospective Studies") | Expanders - Apply equivalent subjects | Interface - EBSCOhost Research Databases | 547,643 |
|  |  | Search modes - Proximity | Search Screen - Advanced Search |  |
|  |  |  | Database - CINAHL Complete |  |
| S14 | S1 OR S2 OR S3 OR S4 OR S5 OR S6 OR S7 OR S8 OR S9 OR S10 OR S11 OR S12 OR S13 | Expanders - Apply equivalent subjects | Interface - EBSCOhost Research Databases | 228,994 |
|  |  | Search modes - Proximity | Search Screen - Advanced Search |  |
|  |  |  | Database - CINAHL Complete |  |
| S13 | "oral health*" OR "dental health*" OR "oral disease*" OR "dental disease*" OR "dental caries*" OR "tooth decay*" OR "oral trauma*" OR "gum disease*" OR periodont* OR gingiv* OR "oral hygiene*" OR "mouth diseas*" OR "dental car*" OR "mouth tumo*r" OR "Mouth Neoplasm*" OR "mouth cancer*" OR "Oral cancer*" OR "Oral traum*" OR "dental visit*" OR "tooth loss*" OR edent* OR "dental status*" OR tooth* OR "dental pain*" OR dentition* OR dental* OR malocclusion* OR teeth* OR xerostomia* OR "tooth diseas*" OR "dental care*" OR "Tooth Injur*" OR toothache* | Expanders - Apply equivalent subjects | Interface - EBSCOhost Research Databases | 201,521 |
|  |  | Search modes - Proximity | Search Screen - Advanced Search |  |
|  |  |  | Database - CINAHL Complete |  |
| S12 | (MH "Tooth Diseases+") OR (MH "Tooth Avulsion") OR (MH "Tooth Abnormalities+") OR (MH "Tooth Injuries+") OR (MH "Tooth, Deciduous+") OR (MH "Tooth Fractures") | Expanders - Apply equivalent subjects | Interface - EBSCOhost Research Databases | 41,049 |
|  |  | Search modes - Proximity | Search Screen - Advanced Search |  |
|  |  |  | Database - CINAHL Complete |  |
| S11 | (MH "Toothache") | Expanders - Apply equivalent subjects | Interface - EBSCOhost Research Databases | 1,002 |
|  |  | Search modes - Proximity | Search Screen - Advanced Search |  |
|  |  |  | Database - CINAHL Complete |  |
| S10 | (MH "Malocclusion+") | Expanders - Apply equivalent subjects | Interface - EBSCOhost Research Databases | 5,952 |
|  |  | Search modes - Proximity | Search Screen - Advanced Search |  |
|  |  |  | Database - CINAHL Complete |  |
| S9 | (MH "Xerostomia+") | Expanders - Apply equivalent subjects | Interface - EBSCOhost Research Databases | 5,593 |
|  |  | Search modes - Proximity | Search Screen - Advanced Search |  |
|  |  |  | Database - CINAHL Complete |  |
| S8 | (MH "Dentition+") OR (MH "Dentition, Secondary") OR (MH "Dentition, Mixed") OR (MH "Dentition, Primary") OR (MH "Tooth, Deciduous+") | Expanders - Apply equivalent subjects | Interface - EBSCOhost Research Databases | 33,070 |
|  |  | Search modes - Proximity | Search Screen - Advanced Search |  |
|  |  |  | Database - CINAHL Complete |  |
| S7 | (MH "Chronic Periodontitis") OR (MH "Aggressive Periodontitis") OR (MH "Periodontitis+") OR (MH "Periodontal Cyst") | Expanders - Apply equivalent subjects | Interface - EBSCOhost Research Databases | 10,300 |
|  |  | Search modes - Proximity | Search Screen - Advanced Search |  |
|  |  |  | Database - CINAHL Complete |  |
| S6 | (MH "Tooth Injuries+") | Expanders - Apply equivalent subjects | Interface - EBSCOhost Research Databases | 3,970 |
|  |  | Search modes - Proximity | Search Screen - Advanced Search |  |
|  |  |  | Database - CINAHL Complete |  |
| S5 | (MH "Dental Caries") | Expanders - Apply equivalent subjects | Interface - EBSCOhost Research Databases | 14,547 |
|  |  | Search modes - Proximity | Search Screen - Advanced Search |  |
|  |  |  | Database - CINAHL Complete |  |
| S4 | (MH "Mouth Neoplasms+") | Expanders - Apply equivalent subjects | Interface - EBSCOhost Research Databases | 15,214 |
|  |  | Search modes - Proximity | Search Screen - Advanced Search |  |
|  |  |  | Database - CINAHL Complete |  |
| S3 | (MH "Dental Care+") | Expanders - Apply equivalent subjects | Interface - EBSCOhost Research Databases | 20,436 |
|  |  | Search modes - Proximity | Search Screen - Advanced Search |  |
|  |  |  | Database - CINAHL Complete |  |
| S2 | (MH "Mouth Diseases+") | Expanders - Apply equivalent subjects | Interface - EBSCOhost Research Databases | 72,205 |
|  |  | Search modes - Proximity | Search Screen - Advanced Search |  |
|  |  |  | Database - CINAHL Complete |  |
| S1 | (MH "Oral Health") | Expanders - Apply equivalent subjects | Interface - EBSCOhost Research Databases | 17,377  Bottom of Form |
|  |  | Search modes - Proximity | Search Screen - Advanced Search |  |
|  |  |  | Database - CINAHL Complete |  |

Embase Classic+Embase <1947 to 2025 May 16>

1	mouth disease/ or mouth injury/ or mouth lesion/ or mouth tumor/ or mouth ulcer/ or tooth disease/ or exp mouth tumor/ or mouth cancer/ or dental caries/ or tooth injury/ or tooth disease/ or periodontitis/ or mouth inflammation/ or periodontal disease/ or dentition/ or View results by resource xerostomia/ or angle class III malocclusion/ or malocclusion/ or angle class II malocclusion/ or angle class I malocclusion/ or tooth pain/ or exp dmf index/	433001

2	(oral health* or dental health* or oral disease* or dental disease* or dental caries* or tooth decay* or oral trauma* or gum disease* or periodont* or gingiv* or oral hygiene* or mouth diseas* or dental car* or mouth tumo?r or Mouth Neoplasm* or mouth cancer* or Oral cancer* or Oral traum* or dental visit* or tooth loss* or edent* or dental status* or tooth* or dental pain* or dentition* or dental* or malocclusion* or teeth* or xerostomia* or tooth diseas* or dental care* or Tooth Injur* or toothache*).mp.	980145

3	1 or 2	1090552

4	cohort analysis/ or observational study/ or follow up/ or longitudinal study/ or prospective study/ or retrospective study/ or observational study/ or observational method/	5319848

5	(Cohort* or follow up* or follow-up* or longitudinal* or prospective* or retrospective* or observational*).mp.	7202461

6	4 or 5	7202461

7	exp Australia/ or New Zealand/	279678

8	(Australia* or oceani* or "New South Wales" or Sydney or Queensland or Brisbane or Victoria or Melbourne or "Australian Capital Territory" or Canberra or "Northern Territory" or Darwin or "South Australia" or Adelaide or Tasmania or Hobart or "Western Australia" or Perth or "New Zealand" or Christchurch or Dunedin or wellington* or Auckland).mp.	505140

9	7 or 8	505140

10	3 and 6 and 9	2208

11	limit 10 to yr="2023 -Current"	**356**

Scopus

**284 results**

( TITLE-ABS-KEY ( "oral health*" OR "dental health*" OR "oral disease*" OR "dental disease*" OR "dental caries*" OR "tooth decay*" OR "oral trauma*" OR "gum disease*" OR periodont* OR gingiv* OR "oral hygiene*" OR "mouth diseas*" OR "dental car*" OR "mouth tumo*r" OR "Mouth Neoplasm*" OR "mouth cancer*" OR "Oral cancer*" OR "Oral traum*" OR "dental visit*" OR "tooth loss*" OR edent* OR "dental status*" OR tooth* OR "dental pain*" OR dentition* OR dental* OR malocclusion* OR teeth* OR xerostomia* OR "tooth diseas*" OR "dental care*" OR "Tooth Injur*" OR toothache* ) AND TITLE-ABS-KEY ( cohort* OR "follow up*" OR follow-up* OR longitudinal* OR prospective* OR retrospective* OR observational* ) AND TITLE-ABS-KEY ( australia* OR oceani* OR "New South Wales" OR sydney OR queensland OR brisbane OR victoria OR melbourne OR "Australian Capital Territory" OR canberra OR "Northern Territory" OR darwin OR "South Australia" OR adelaide OR tasmania OR hobart OR "Western Australia" OR perth OR "New Zealand" OR christchurch OR dunedin OR wellington* OR auckland ) ) AND PUBYEAR > 2022 AND PUBYEAR < 2026

**Web of Science**

| # | Search Query | Database | Results |
| --- | --- | --- | --- |
| 1 | australia* OR oceani* OR "New South Wales" OR sydney OR queensland OR brisbane OR victoria OR melbourne OR "Australian Capital Territory" OR canberra OR "Northern Territory" OR darwin OR "South Australia" OR adelaide OR tasmania OR hobart OR "Western Australia" OR perth OR "New Zealand" OR christchurch OR dunedin OR wellington* OR auckland (Topic) | Web of Science Core Collection | 854418 |
| 2 | cohort* OR "follow up*" OR follow-up* OR longitudinal* OR prospective* OR retrospective* OR observational* (Topic) | Web of Science Core Collection | 4617639 |
| 3 | "oral health*" OR "dental health*" OR "oral disease*" OR "dental disease*" OR "dental caries*" OR "tooth decay*" OR "oral trauma*" OR "gum disease*" OR periodont* OR gingiv* OR "oral hygiene*" OR "mouth diseas*" OR "dental car*" OR "mouth tumo*r" OR "Mouth Neoplasm*" OR "mouth cancer*" OR "Oral cancer*" OR "Oral traum*" OR "dental visit*" OR "tooth loss*" OR edent* OR "dental status*" OR tooth* OR "dental pain*" OR dentition* OR dental* OR malocclusion* OR teeth* OR xerostomia* OR "tooth diseas*" OR "dental care*" OR "Tooth Injur*" OR toothache* (Topic) | Web of Science Core Collection | 610662 |
| 4 | #1 AND #2 AND #3 | Web of Science Core Collection | 1155 |
| 5 | #1 AND #2 AND #3 and 2023 or 2024 or 2025 (Publication Years) | Web of Science Core Collection | **174** |
