## Appendix 3 for "Oral Health Research Across the Lifespan: A Systematic Mapping Review of Cohort Studies in Australia and New Zealand"

Maternal alcohol disorder and dental hospital admissions: Data-linkage to determine potential pathways

Slack-Smith, L. M.; O'Leary, C.

*May be an abstract

*Couldn’t find it in Google Search as well

Excluded: it is an Abstract

Oral cavity cancer outcomes at a tertiary hospital: Sir charles gairdner hospital, Perth, Australia

DOI: [10.1177/0194599818787193d](https://dx.doi.org/10.1177/0194599818787193d)

Hendriks, T.; Cardemil, F.

*An abstract

Excluded: it is an Abstract

The Australian Aboriginal Birth Cohort Study, 1987 2007: Is birth size a predictor of oral health outcomes?

Jamieson, L.

DOI: [10.1016/s0378-3782(07)70325-0](https://dx.doi.org/10.1016/s0378-3782(07)70325-0)

*Poster

Excluded: it is a poster

Is sustained breastfeeding associated with early childhood caries?

Devenish, G.; Scott, J.; Begley, A.; Spencer, J.; Thomson, M.; Ha, D.; Do, L.

DOI: [10.1111/mcn.12933](https://dx.doi.org/10.1111/mcn.12933)

*An abstract

Excluded: it is an Abstract

Dietary intake of nutrients and compromised periodontal health: The concord health and ageing men project

Milledge, K.; Cumming, R.; Wright, F.; Naganathan, V.; Blyth, F.; Le Couteur, D.; Hirani, V.

Annals of Nutrition and Metabolism 2017;71(Supplement 2)():359-360

2017

DOI: [10.1159/000480486](https://dx.doi.org/10.1159/000480486)

*An abstract

Excluded: it is an Abstract

Further evidence that periodontal bone loss increases with smoking and age

Zeng J, Williams SM, Fletcher DJ, Cameron CM, Broadbent JM, Shearer DM, Thomson WM.

Evidence-Based Dentistry (2014) 15, 72-73. doi:10.1038/sj.ebd.6401038

*Commentary

Excluded: it is a commentary

Smoking cessation and periodontal health – a missed opportunity?

Thomson WM, Broadbent JM, Welch D, Beck JD, Poulton R.

Evidence-Based Dentistry (2009) 10, 18-19. doi:10.1038/sj.ebd.6400632

*Commentary

Excluded: it is a commentary

Periodontitis and risk of all-cause and cardiovascular mortality in adults with end-stage kidney disease: A propensity-matched study

Palmer, S.; Ruospo, M.; Wong, G.; Craig, J.; Johnson, D. W.; Ford, P.; Tonelli, M.; Petruzzi, M.; De Benedittis, M.; Strippoli, G.

Nephrology September 2015;3)():51 2015 September

Excluded: this study has been conducted outside of Australia/New Zealand

Does orthodontic treatment in early adolescence positively influence psychosocial wellbeing in adulthood?

Madurantakam, P. Evidence-Based Dentistry 12 2019;20(4):107-108 2019 12

Excluded: it is a commentary paper

Summary of: Permanent dentition caries through the half of life

Broadbent, J. M.; Page, L. A. F.; Thomson, W. M.; Poulton, R.

British Dental Journal Oct 2013;215(7):342-343 2013 Oct

Excluded: it is a commentary paper

The effectiveness of a sustained nurse home visiting intervention for Aboriginal infants compared with non-Aboriginal infants and with Aboriginal infants receiving usual child health care: a quasi-experimental trial - the Bulundidi Gudaga study

Kemp, L.; Grace, R.; Comino, E.; Jackson Pulver, L.; McMahon, C.; Harris, E.; Harris, M.; George, A.; Mack, H. A.

BMC Health Services Research 08 03 2018;18(1):599 2018 08 03

Excluded: it is not a cohort study

Associations between discrimination and dental visiting behaviours in an Aboriginal Australian birth cohort

Jamieson, L. M.; Steffens, M.; Paradies, Y. C.

Australian and New Zealand Journal of Public Health Feb 2013;37(1)():92-93

2013 Feb

DOI: 10.1111/1753-6405.12018

Excluded: it is a letter
