## Appendix 5 for "Oral Health Research Across the Lifespan: A Systematic Mapping Review of Cohort Studies in Australia and New Zealand"

**Cohort Studies with Primary Data Collections**

| # | Study Name | Publication APA Citations |
| --- | --- | --- |
| 1 | **Australian Research Centre for Population Oral Health Cohort Study—The University of Adelaide** | **Allister, J. H., Spencer, A. J., & Brennan, D. S. (1996). Provision of orthodontic care to adolescents in South Australia: The type, the provider, and the place of treatment. Australian Dental Journal, 41(6), 405–410. https://doi.org/10.1111/j.1834-7819.1996.tb06027.x**  **Arrow, P., Brennan, D. S., & Spencer, A. J. (2012). Social acceptability of dental appearance and benefits of fixed orthodontic treatment: A 17-year observational cohort study. Journal of Public Health Dentistry, 72(2), 135–142. https://doi.org/10.1111/j.1752-7325.2011.00293.x**  **Brennan, D. S., & Spencer, A. J. (2014). Childhood oral health and SES predictors of caries in 30-year-olds. Caries Research, 48(3), 237–243. https://doi.org/10.1159/000354044**  **Doğramacı, E. J., & Brennan, D. S. (2019a). The influence of orthodontic treatment on dental caries: An Australian cohort study. Community Dentistry and Oral Epidemiology, 47(3), 210–216. https://doi.org/10.1111/cdoe.12446**  **Doğramacı, E. J., & Brennan, D. S. (2019b). The long-term influence of orthodontic treatment on adults' psychosocial outcomes: An Australian cohort study. Orthodontics & Craniofacial Research, 22(4), 312–320. https://doi.org/10.1111/ocr.12327**  **Spencer, A. J., Allister, J. H., & Brennan, D. S. (1995). Predictors of fixed orthodontic treatment in 15-year-old adolescents in South Australia. Community Dentistry and Oral Epidemiology, 23(6), 350–355. https://doi.org/10.1111/j.1600-0528.1995.tb00261.x** |
| 2 | **Mater Mother’s Hospital Study_2003** | **Wan, A. K. L., Seow, W. K., Purdie, D. M., Bird, P. S., Walsh, L. J., & Tudehope, D. I. (2003). A longitudinal study of Streptococcus mutans colonization in infants after tooth eruption. Journal of Dental Research, 82(7), 504–508. https://doi.org/10.1177/154405910308200703**  **Wan, A. K. L., Seow, W. K., Purdie, D. M., Bird, P. S., Walsh, L. J., & Tudehope, D. I. (2003). Immunoglobulins in saliva of preterm and full-term infants: A longitudinal study from 0–18 months of age. Oral Microbiology and Immunology, 18(2), 72–78. https://doi.org/10.1034/j.1399-302x.2003.00016.x** |
| 3 | **Aboriginal Birth Cohort Study (ABC)** | **Jamieson, L. M., Armfield, J. M., Roberts-Thomson, K. F., & Sayers, S. M. (2010). A retrospective longitudinal study of caries development in an Australian Aboriginal birth cohort. Caries Research, 44(4), 415–420. https://doi.org/10.1159/000316665**  **Jamieson, L. M., Do, L. G., Bailie, R. S., Sayers, S. M., & Turrell, G. (2013). Associations between area-level disadvantage and DMFT among a birth cohort of Indigenous Australians. Australian Dental Journal, 58(1), 75–81. https://doi.org/10.1111/adj.12017**  **Jamieson, L. M., Gunthorpe, W., Cairney, S. J., Sayers, S. M., Roberts-Thomson, K. F., & Slade, G. D. (2010). Substance use and periodontal disease among Australian Aboriginal young adults. Addiction, 105(4), 719–726. https://doi.org/10.1111/j.1360-0443.2009.02851.x**  **Jamieson, L. M., Paradies, Y. C., Gunthorpe, W., Cairney, S. J., & Sayers, S. M. (2011). Oral health and social and emotional well-being in a birth cohort of Aboriginal Australian young adults. BMC Public Health, 11, Article 656. https://doi.org/10.1186/1471-2458-11-656**  **Jamieson, L. M., Roberts-Thomson, K. F., & Sayers, S. M. (2010). Dental caries risk indicators among Australian Aboriginal young adults. Community Dentistry and Oral Epidemiology, 38(3), 213–221. https://doi.org/10.1111/j.1600-0528.2009.00519.x**  **Jamieson, L. M., Roberts-Thomson, K. F., & Sayers, S. M. (2010). Risk indicators for severe impaired oral health among indigenous Australian young adults. BMC Oral Health, 10, Article 1. https://doi.org/10.1186/1472-6831-10-1**  **Jamieson, L. M., & Sayers, S. M. (2008). Oral health investigations of indigenous participants in remote settings: A methods paper describing the dental component of wave III of an Australian Aboriginal birth cohort study. BMC Oral Health, 8, Article 24. https://doi.org/10.1186/1472-6831-8-24**  **Jamieson, L. M., Sayers, S. M., & Roberts-Thomson, K. F. (2010). Clinical oral health outcomes in young Australian Aboriginal adults compared with national-level counterparts. Medical Journal of Australia, 192(10), 558–561. https://doi.org/10.5694/j.1326-5377.2010.tb03635.x**  **Jamieson, L. M., Sayers, S. M., & Roberts-Thomson, K. F. (2013). Associations between oral health and height in an Indigenous Australian birth cohort. Community Dental Health, 30(1), 58–64.**  **Sayers, S. M., Mackerras, D., & Singh, G. R. (2017). Cohort Profile: The Australian Aboriginal Birth Cohort (ABC) study. International Journal of Epidemiology, 46(5), 1383–1383f. https://doi.org/10.1093/ije/dyw291** |
| 4 | **University of Adelaide Twin Study (UAT)** | **Bockmann, M. R., Harris, A. V., Bennett, C. N., Odeh, R., Hughes, T. E., & Townsend, G. C. (2011). Timing of colonization of caries-producing bacteria: An approach based on studying monozygotic twin pairs. International Journal of Dentistry, 2011, Article 571573. https://doi.org/10.1155/2011/571573**  **Chan, E., Bockmann, M., Hughes, T., Mihailidis, S., & Townsend, G. (2012). Do feeding practices, gestation length, and birth weight affect the timing of emergence of the first primary tooth? In G. Townsend, E. Kanazawa, & H. Takayama (Eds.), New directions in dental anthropology: Paradigms, methodologies and outcomes (pp. 35–45). University of Adelaide Press.**  **Corruccini, R. S., Townsend, G. C., & Schwerdt, W. (2005). Correspondence between enamel hypoplasia and odontometric bilateral asymmetry in Australian twins. American Journal of Physical Anthropology, 126(2), 177–182. https://doi.org/10.1002/ajpa.20113**  **Hughes, T., Bockmann, M., Mihailidis, S., Bennett, C., Harris, A., Seow, W. K., Lekkas, D., Ranjitkar, S., Rupinskas, L., Pinkerton, S., Brook, A., Smith, R., & Townsend, G. C. (2013). Genetic, epigenetic, and environmental influences on dentofacial structures and oral health: Ongoing studies of Australian twins and their families. Twin Research and Human Genetics, 16(1), 43–51. https://doi.org/10.1017/thg.2012.78**  **Paul, K. S., Stojanowski, C. M., Hughes, T., Brook, A., & Townsend, G. C. (2021). The genetic architecture of anterior tooth morphology in a longitudinal sample of Australian twins and families. Archives of Oral Biology, 129, 105168. https://doi.org/10.1016/j.archoralbio.2021.105168**  **Paul, K. S., Stojanowski, C. M., Hughes, T., Brook, A. H., & Townsend, G. C. (2022). Genetic correlation, pleiotropy, and molar morphology in a longitudinal sample of Australian twins and families. Genes, 13(6), 996. https://doi.org/10.3390/genes13060996**  **Race, J. P., Townsend, G. C., & Hughes, T. E. (2006). Chorion type, birthweight discordance and tooth-size variability in Australian monozygotic twins. Twin Research and Human Genetics, 9(2), 285–291. https://doi.org/10.1375/twin.9.2.285**  **Taduran, R. J. O., Ranjitkar, S., Hughes, T., Townsend, G., & Brook, A. H. (2016). Complex systems in human development: Sexual dimorphism in teeth and fingerprints of Australian twins. International Journal of Design & Nature and Ecodynamics, 11(4), 676–685. https://doi.org/10.2495/DNE-V11-N4-676-685**  **Taji, S. S., Seow, W. K., Townsend, G. C., & Holcombe, T. (2010). A controlled study of dental erosion in 2- to 4-year-old twins. International Journal of Paediatric Dentistry, 20(6), 400–409. https://doi.org/10.1111/j.1365-263X.2010.01081.x**  **Townsend, G., Richards, L., Messer, L. B., Hughes, T., Pinkerton, S., Seow, K., Gotjamanos, T., Gully, N., & Bockmann, M. (2006). Genetic and environmental influences on dentofacial structures and oral health: Studies of Australian twins and their families. Twin Research and Human Genetics, 9(6), 727–732.** [**https://doi.org/10.1375/twin.9.6.727**](https://doi.org/10.1375/twin.9.6.727)  **Apps, M. V. B., Hughes, T. E., & Townsend, G. C. (2004). The effect of birthweight on tooth-size variability in twins. Twin Research, 7(5), 415–420. https://doi.org/10.1375/twin.7.5.415**  **Brown, T., Townsend, G. C., Richards, L. C., & Travan, G. R. (1987). A study of dentofacial morphology in South Australian twins. Australian Dental Journal, 32(2), 81–90.**  **Dempsey, P. J., & Townsend, G. C. (2001). Genetic and environmental contributions to variation in human tooth size. Heredity, 86, 685–693.**  **Hughes, T. E., Bockmann, M. R., Seow, K., Gotjamanos, T., Gully, N., Richards, L. C., & Townsend, G. C. (2007). Strong genetic control of emergence of human primary incisors. Journal of Dental Research, 86(12), 1160–1165.**  **Hughes, T. E., Townsend, G. C., Pinkerton, S. K., Bockmann, M. R., Seow, W. K., Brook, A. H., Richards, L. C., Mihailidis, S., Ranjitkar, S., & Lekkas, D. (2014). The teeth and faces of twins: providing insights into dentofacial development and oral health for practising oral health professionals. Australian Dental Journal, 59(1 Suppl), 101–116. https://doi.org/10.1111/adj.12101**  **Ooi, G., Townsend, G., & Seow, W. K. (2013). Bacterial colonization, enamel defects and dental caries in 4–6-year-old mono- and dizygotic twins. International Journal of Paediatric Dentistry. https://doi.org/10.1111/ipd.12041**  **Tadros, M., Brook, A. H., Ranjitkar, S., & Townsend, G. C. (2019). Compensatory interactions between developing maxillary anterior teeth in a sample of twins. Archives of Oral Biology, 97, 198–207. https://doi.org/10.1016/j.archoralbio.2018.10.010**  **Taduran, R. J. O., Ranjitkar, S., Hughes, T., Townsend, G., & Brook, A. H. (2016). Complex systems in human development: Sexual dimorphism in teeth and fingerprints of Australian twins. International Journal of Design & Nature and Ecodynamics, 11(4), 676–685. https://doi.org/10.2495/DNE-V11-N4-676-685**  **Taji, S. S., Seow, W. K., Townsend, G. C., & Holcombe, T. (2011). Enamel hypoplasia in the primary dentition of monozygotic and dizygotic twins compared with singleton controls. International Journal of Paediatric Dentistry, 21(3), 175–184. https://doi.org/10.1111/j.1365-263X.2010.01106.x** |
| 5 | **Christchurch Child Development Study** | **Beautrais, A. L., Fergusson, D. M., & Shannon, F. T. (1982). Use of preschool dental services in a New Zealand birth cohort. Community Dentistry and Oral Epidemiology, 10, 249–252.**  **Fergusson, D. M., & Horwood, L. J. (1986). Relationships between exposure to additional fluoride, social background and dental health in 7-year-old children. Community Dentistry and Oral Epidemiology, 14, 48–52.**  **Ruiz, B., Broadbent, J. M., Thomson, W. M., Ramrakha, S., Boden, J., Horwood, J., & Poulton, R. (2023). Is childhood oral health the ‘canary in the coal mine’ for poor adult general health? Findings from two New Zealand birth cohort studies. Community Dentistry and Oral Epidemiology, 51, 838–846. https://doi.org/10.1111/cdoe.12772** |
| 6 | **Concord Health and Ageing in Men Project (CHAMP)** | **Beautrais, A. L., Fergusson, D. M., & Shannon, F. T. (1982). Use of preschool dental services in a New Zealand birth cohort. Community Dentistry and Oral Epidemiology, 10, 249–252.**  **Fergusson, D. M., & Horwood, L. J. (1986). Relationships between exposure to additional fluoride, social background and dental health in 7-year-old children. Community Dentistry and Oral Epidemiology, 14, 48–52.**  **Ruiz, B., Broadbent, J. M., Thomson, W. M., Ramrakha, S., Boden, J., Horwood, J., & Poulton, R. (2023). Is childhood oral health the ‘canary in the coal mine’ for poor adult general health? Findings from two New Zealand birth cohort studies. Community Dentistry and Oral Epidemiology, 51, 838–846. https://doi.org/10.1111/cdoe.12772**  **Takehara, S., Hirani, V., Wright, F. A. C., Naganathan, V., Blyth, F. M., Le Couteur, D. G., Waite, L. M., Seibel, M. J., Handelsman, D. J., & Cumming, R. G. (2021). Appetite, oral health and weight loss in community-dwelling older men: An observational study from the Concord Health and Ageing in Men Project (CHAMP). BMC Geriatrics, 21(1), Article 255. https://doi.org/10.1186/s12877-021-02169-y**  **Takehara, S., Wright, F. A. C., Waite, L. M., Naganathan, V., Hirani, V., Blyth, F. M., Le Couteur, D. G., Seibel, M. J., Handelsman, D. J., & Cumming, R. G. (2020). Oral health and cognitive status in the Concord Health and Ageing in Men Project: A cross-sectional study in community-dwelling older Australian men. Gerodontology, 1–8. https://doi.org/10.1111/ger.12469**  **Tran, J., Wright, F. A. C., Takara, S., Shu, C.-C., Chu, S. K.-Y., Naganathan, V., Hirani, V., Blyth, F. M., Le Couteur, D. G., Waite, L. M., Handelsman, D. J., Seibel, M. J., Milledge, K. L., & Cumming, R. G. (2019). Oral health behaviours of older Australian men: The Concord Health and Ageing in Men Project. Australian Dental Journal, 64, 246–255. https://doi.org/10.1111/adj.12694**  **Wright, F. A. C., Chu, S. K.-Y., Milledge, K. L., Valdez, E., Law, G., Hsu, B., Naganathan, V., Hirani, V., Blyth, F. M., Le Couteur, D. G., Harford, J., Waite, L. M., Handelsman, D. J., Seibel, M. J., & Cumming, R. G. (2018). Oral health of community-dwelling older Australian men: The Concord Health and Ageing in Men Project (CHAMP). Australian Dental Journal, 63, 55–65. https://doi.org/10.1111/adj.12564**  **Wright, F. A. C., Shu, E. C.-C., Cumming, R. G., Naganathan, V., Blyth, F. M., Hirani, V., Le Couteur, D. G., Handelsman, D. J., Seibel, M. J., Waite, L. M., & Stanaway, F. F. (2023). Oral health-related quality of life of older Australian men. Community Dentistry and Oral Epidemiology, 51, 767–777. https://doi.org/10.1111/cdoe.12754** |
| 7 | **Dunedin Multidisciplinary Health and Development Study (DMHDS)** | **Broadbent, J. M., Thomson, W. M., Boyens, J. V., & Poulton, R. (2011). Dental plaque and oral health during the first 32 years of life. The Journal of the American Dental Association, 142(4), 415–426. https://doi.org/10.14219/jada.archive.2011.0192**  **Broadbent, J. M., Thomson, W. M., & Williams, S. M. (2005). Does caries in primary teeth predict enamel defects in permanent teeth? A longitudinal study. Journal of Dental Research, 84(3), 260–264. https://doi.org/10.1177/154405910508400311**  **Broadbent, J. M., Williams, K. B., Thomson, W. M., & Williams, S. M. (2006). Dental restorations: a risk factor for periodontal attachment loss? Journal of Clinical Periodontology, 33(11), 803–810. https://doi.org/10.1111/j.1600-051x.2006.00988.x**  **Morelli, E. L., Broadbent, J. M., Knight, E. T., Leichter, J. W., & Thomson, W. M. (2022). Does having children affect women’s oral health? A longitudinal study. Journal of Public Health Dentistry, 82(1), 31–39. https://doi.org/10.1111/jphd.12466**  **Olliver, S. J., Broadbent, J. M., Prasad, S., Cai, C., Thomson, W. M., & Farella, M. (2022). Changes in incisor relationship over the life course - Findings from a cohort study. Journal of Dentistry, 117, 103919. https://doi.org/10.1016/j.jdent.2021.103919**  **Ruiz, B., Broadbent, J. M., Thomson, W. M., Ramrakha, S., Hong, C. L., & Poulton, R. (2023). Differential unmet needs and experience of restorative dental care in trajectories of dental caries experience: A birth cohort study. Caries Research, 57(4), 524–535. https://doi.org/10.1159/000530378**  **Shearer, D. M., Thomson, W. M., Broadbent, J. M., & Poulton, R. (2011). Does maternal oral health predict child oral health-related quality of life in adulthood? Health and Quality of Life Outcomes, 9(1), 50. https://doi.org/10.1186/1477-7525-9-50**  **Thomson, W. M., Broadbent, J. M., Poulton, R., & Beck, J. D. (2006). Changes in periodontal disease experience from 26 to 32 years of age in a birth cohort. Journal of Periodontology, 77(6), 947–954. https://doi.org/10.1902/jop.2006.050319**  **Thomson, W. M., Broadbent, J. M., Welch, D., Beck, J. D., & Poulton, R. (2007). Cigarette smoking and periodontal disease among 32-year-olds: A prospective study of a representative birth cohort. Journal of Clinical Periodontology, 34(10), 828–834. https://doi.org/10.1111/j.1600-051x.2007.01131.x**  **Thomson, W. M., Poulton, R., Broadbent, J. M., Moffitt, T. E., Caspi, A., Beck, J. D., Welch, D., & Hancox, R. J. (2008). Cannabis smoking and periodontal disease among young adults. JAMA, 299(5), 525–531.** [**https://doi.org/10.1001/jama.299.5.525**](https://doi.org/10.1001/jama.299.5.525)  **Broadbent, J. M., & Thomson, W. M. (2008). Long-term survival of enamel defect-affected teeth. Journal of Dental Research, 87(3), 288–292. https://doi.org/10.1177/154405910808700302**  **Broadbent, J. M., Thomson, W. M., & Poulton, R. (2006). Oral health-related beliefs, behaviors, and outcomes through the life course. Community Dentistry and Oral Epidemiology, 34(5), 348–358. https://doi.org/10.1111/j.1600-0528.2006.00293.x**  **Broadbent, J. M., Thomson, W. M., & Poulton, R. (2006). Permanent dentition caries through the first half of life: a birth cohort study. Caries Research, 40(6), 459–465. https://doi.org/10.1159/000095641**  **Broadbent, J. M., Thomson, W. M., & Poulton, R. (2009). Impact of dental visiting trajectory patterns on clinical oral health and oral health-related quality of life. Journal of Public Health Dentistry, 69(3), 128–132. https://doi.org/10.1111/j.1752-7325.2009.00115.x**  **Broadbent, J. M., Thomson, W. M., Williams, S. M., & Poulton, R. (2008). Is Asthma a Risk Factor for Dental Caries? Findings from a Cohort Study. Caries Research, 42(4), 265–270. https://doi.org/10.1159/000135639**  **Lawrence, H. P., Thomson, W. M., Broadbent, J. M., & Poulton, R. (2008). Oral health-related quality of life in a birth cohort of 32-year olds. Community Dentistry and Oral Epidemiology, 36(4), 305–316. https://doi.org/10.1111/j.1600-0528.2007.00404.x**  **Leichter, J. W., Broadbent, J. M., Thomson, W. M., Williams, M. J. A., Farella, M., & Poulton, R. (2021). Periodontitis and multiple markers of cardiometabolic risk in the fourth decade: A prospective study of a New Zealand birth cohort. Journal of Periodontology, 92(2), 233–241. https://doi.org/10.1002/JPER.19-0657**  **Shearer, D. M., Thomson, W. M., Broadbent, J. M., & Poulton, R. (2011). Maternal oral health predicts their children’s caries experience in adulthood. Journal of Dental Research, 90(5), 672–677. https://doi.org/10.1177/0022034510393354**  **Thomson, W. M., Poulton, R., & Kruger, E. (2000). Family history of oral health: Findings from the Dunedin Study. New Zealand Dental Journal, 96(424), 48–51.**  **Thomson, W. M., Williams, S. M., Broadbent, J. M., Poulton, R., & Locker, D. (2010). Long-term dental visiting patterns and adult oral health. Journal of Dental Research, 89(3), 307–311.** [**https://doi.org/10.1177/0022034509356779**](https://doi.org/10.1177/0022034509356779)  **Broadbent, J. M., Thomson, W. M., & Poulton, R. (2006). Progression of dental caries and tooth loss between the third and fourth decades of life: A birth cohort study. Caries Research, 40(6), 459–465. https://doi.org/10.1159/000095641**  **Broadbent, J. M., Thomson, W. M., & Poulton, R. (2007). Reexamining the association between smoking and periodontitis in the Dunedin study. The Journal of the American Dental Association, 138(11), 1432–1433. https://doi.org/10.14219/jada.archive.2007.0082**  **Broadbent, J. M., Thomson, W. M., & Poulton, R. (2008). Trajectory patterns of dental caries experience in the permanent dentition to the fourth decade of life. Journal of Dental Research, 87(1), 69–72. https://doi.org/10.1177/154405910808700113**  **Hashim, R., Thomson, W. M., & Pack, A. R. C. (2001). Smoking in adolescence as a predictor of early loss of periodontal attachment. Community Dentistry and Oral Epidemiology, 29(2), 130–135. https://doi.org/10.1034/j.1600-0528.2001.029002130.x**  **Poulton, R., Caspi, A., Milne, B. J., Thomson, W. M., Taylor, A., Sears, M. R., & Moffitt, T. E. (2002). Association between children's experience of socioeconomic disadvantage and adult health: a life-course study. The Lancet, 360(9346), 1640–1645. https://doi.org/10.1016/s0140-6736(02)11602-3**  **Thomson, W. M., Caspi, A., Poulton, R., Moffitt, T. E., & Broadbent, J. M. (2011). Personality and oral health. European Journal of Oral Sciences, 119(5), 366–372. https://doi.org/10.1111/j.1600-0722.2011.00845.x**  **Thomson, W. M., Hashim, R., & Pack, A. R. C. (2000). The prevalence and intraoral distribution of periodontal attachment loss in a birth cohort of 26-year-olds. Journal of Periodontology, 71(12), 1840–1845. https://doi.org/10.1902/jop.2000.71.12.1840**  **Thomson, W. M., Poulton, R., & Kruger, E. (2000). Family history of oral health: Findings from the Dunedin Study. New Zealand Dental Journal, 96(424), 48–51.**  **Thomson, W. M., Poulton, R., Kruger, E., & Boyd, D. (2000). Socio-economic and behavioural risk factors for tooth loss from age 18 to 26 among participants in the Dunedin Multidisciplinary Health and Development Study. Caries Research, 34(5), 361–366. https://doi.org/10.1159/000016611**  **Thomson, W. M., Shearer, D. M., Broadbent, J. M., Foster Page, L. A., & Poulton, R. (2013). The natural history of periodontal attachment loss during the third and fourth decades of life. Journal of Clinical Periodontology, 40(7), 672–680.** [**https://doi.org/10.1111/jcpe.12105**](https://doi.org/10.1111/jcpe.12105)  **Broadbent, J. M., Poulton, R., & Thomson, W. M. (2009). Dental visiting trajectory patterns and their antecedents. Journal of Public Health Dentistry, 69(1), 43–48. https://doi.org/10.1111/j.1752-7325.2008.00096.x**  **Cacic, N., Knight, E. T., Broadbent, J. M., Lee, M., & Farella, M. (2021). Occlusal features and TMJ clicking: A 30-year evaluation from a cohort study. Journal of Oral Rehabilitation, 48(10), 1146–1154. https://doi.org/10.1111/joor.13227**  **Cai, C., Farella, M., Eli, A., Broadbent, J. M., & Poulton, R. (2021). Occurrence, associations, and impacts of nocturnal parafunction, daytime parafunction, and temporomandibular symptoms in 38-year-old individuals. The Journal of the American Dental Association, 152(6), 465–474. https://doi.org/10.1016/j.adaj.2020.11.026**  **Gao, X., Broadbent, J. M., & Thomson, W. M. (2019). Periodontitis is not associated with metabolic risk during the fourth decade of life. Journal of Clinical Periodontology, 46(7), 712–720. https://doi.org/10.1111/jcpe.13136**  **Locker, D., Poulton, R., & Thomson, W. M. (2000). Psychological disorder, conditioning experiences, and the onset of dental anxiety in early adulthood. Journal of Dental Research, 79(7), 1585–1590. https://doi.org/10.1177/00220345000790071801**  **Locker, D., Thomson, W. M., & Poulton, R. (2001). Dental fear with and without blood-injection fear: Implications for dental health and clinical practice. Journal of Public Health Dentistry, 61(2), 98–103. https://doi.org/10.1111/j.1752-7325.2001.tb03367.x**  **Meier, M. H., Caspi, A., Cerdá, M., Hancox, R. J., Harrington, H., Houts, R., Poulton, R., Ramrakha, S., Thomson, W. M., & Moffitt, T. E. (2016). Associations between cannabis use and physical health problems in early midlife: A longitudinal comparison of persistent cannabis vs tobacco users. JAMA Psychiatry, 73(7), 731–740. https://doi.org/10.1001/jamapsychiatry.2016.0637**  **Poulton, R., Thomson, W. M., Davies, S., Kruger, E., Brown, R. H., & Silva, P. A. (1997). Good teeth, bad teeth and fear of the dentist. Behaviour Research and Therapy, 35(4), 327–334. https://doi.org/10.1016/s0005-7967(96)00099-2**  **Poulton, R., Waldie, K. E., Thomson, W. M., & Locker, D. (2001). Determinants of early- vs. late-onset dental fear in a longitudinal-epidemiological study. Behaviour Research and Therapy, 39(7), 777–785. https://doi.org/10.1016/s0005-7967(00)00058-1**  **Thomson, W. M., Poulton, R., & Locker, D. (2000). Inter-generational continuity in periodontal health: findings from the Dunedin family history study. New Zealand Dental Journal, 96(425), 90–93.**  **Broadbent, J. M., Thomson, W. M., Williams, M. J., & Poulton, R. (2021). Antecedents and associations of root surface caries experience among 38-year-olds. Caries Research, 55(2), 137–145. https://doi.org/10.1159/000514101**  **Evans, R. W., Beck, D. J., Brown, R. H., & Silva, P. A. (1984). Relationship between fluoridation and socioeconomic status on dental caries experience in 5-year-old New Zealand children. Community Dentistry and Oral Epidemiology, 12(1), 5–9. https://doi.org/10.1111/j.1600-0528.1984.tb01407.x**  **Hong, C. L., Broadbent, J. M., Thomson, W. M., & Poulton, R. (2020a). The Dunedin Multidisciplinary Health and Development Study: Oral health findings and their implications. Journal of the Royal Society of New Zealand, 50(1), 35–46. https://doi.org/10.1080/03036758.2019.1678516**  **Hong, C. L., Broadbent, J. M., Thomson, W. M., & Poulton, R. (2020b). Validity of self-reported periodontal questions in a New Zealand cohort. Journal of Clinical Periodontology, 47(3), 308–316. https://doi.org/10.1111/jcpe.13247**  **5. Lewsey, J. D., & Thomson, W. M. (2004). The utility of the zero-inflated Poisson and zero-inflated negative binomial models: A case study of cross-sectional and longitudinal DMF data examining the effect of socio-economic status. Community Dentistry and Oral Epidemiology, 32(3), 183–189. https://doi.org/10.1111/j.1600-0528.2004.00142.x**  **Poulton, R., Thomson, W. M., Brown, R. H., & Silva, P. A. (1997). Psychological disorders and dental anxiety in a young adult population. Behaviour Research and Therapy, 35(4), 287–293. https://doi.org/10.1016/s0005-7967(96)00115-8**  **Suckling, G. W., Herbison, G. P., & Brown, R. H. (1987). Etiological factors influencing the prevalence of developmental defects of dental enamel in nine-year-old New Zealand children participating in a health and development study. Journal of Dental Research, 66(9), 1466–1469. https://doi.org/10.1177/00220345870660091301**  **Thomson, W. M. (2002). Orthodontic treatment outcomes in the long term: findings from a longitudinal study of New Zealanders. The Angle Orthodontist, 72(5), 449–455. https://doi.org/10.1043/0003-3219(2002)072&lt;0449:otoitl>2.0.co;2**  **Thomson, W. M., Broadbent, J. M., Caspi, A., Poulton, R., & Moffitt, T. E. (2019). Childhood IQ predicts age-38 oral disease experience and service-use. Community Dentistry and Oral Epidemiology, 47(3), 252–258. https://doi.org/10.1111/cdoe.12450**  **Thomson, W. M., Poulton, R., Kruger, E., & Boyd, D. (2000). Third molar outcomes from age 18 to 26: findings from a population-based New Zealand longitudinal study. Journal of Oral and Maxillofacial Surgery, 58(7), 770–774.** [**https://doi.org/10.1053/joms.2000.7247**](https://doi.org/10.1053/joms.2000.7247)  **Broadbent, J. M., Poulton, R., & Thomson, W. M. (2007). Trajectories of dental anxiety in a birth cohort. Journal of Dental Research, 86(6), 553–556. https://doi.org/10.1177/154405910708600613**  **Broadbent, J. M., Thomson, W. M., Williams, S. M., Poulton, R., & Moffitt, T. E. (2012). Telomere length and periodontal attachment loss: A prospective cohort study. Journal of Dental Research, 91(1), 41–45. https://doi.org/10.1177/0022034511425121**  **Locker, D., Liddell, A., & Shapiro, D. (1999). Incidence of dental anxiety in young adults in relation to dental treatment experience. Community Dentistry and Oral Epidemiology, 27(5), 368–373. https://doi.org/10.1111/j.1600-0528.1999.tb02035.x**  **Locker, D., Poulton, R., & Thomson, W. M. (2001). Changes in self-reported dental anxiety in New Zealand adolescents from ages 15 to 18 years. International Journal of Paediatric Dentistry, 11(4), 245–251. https://doi.org/10.1046/j.1365-263x.2001.00287.x**  **Thomson, W. M. (2012). Social inequality in oral health. Community Dentistry and Oral Epidemiology, 40(Suppl. 2), 28–32. https://doi.org/10.1111/j.1600-0528.2012.00718.x**  **Thomson, W. M., Edwards, S. J., Dobson-Le, D. P., & Poulton, R. (2005). IL-1 genotype and adult periodontitis among young New Zealanders. Journal of Dental Research, 84(1), 52–55. https://doi.org/10.1177/154405910508400108**  **Thomson, W. M., Lawrence, H. P., Broadbent, J. M., & Poulton, R. (2006). The impact of xerostomia on oral-health-related quality of life among younger adults. Health and Quality of Life Outcomes, 4, Article 86. https://doi.org/10.1186/1477-7525-4-86**  **Thomson, W. M., & Locker, D. (2000). Dental neglect and dental health among 26-year-olds in the Dunedin Multidisciplinary Health and Development Study. Community Dentistry and Oral Epidemiology, 28(6), 414–418. https://doi.org/10.1034/j.1600-0528.2000.028006414.x**  **Thomson, W. M., Mejia, G. C., Broadbent, J. M., & Poulton, R. (2012). Construct validity of Locker’s global oral health item. Journal of Dental Research, 91(11), 1038–1042.** [**https://doi.org/10.1177/0022034512460830**](https://doi.org/10.1177/0022034512460830)  **Barker, M. J., Thomson, W. M., & Poulton, R. (2005). Personality traits in adolescence and satisfaction with orthodontic treatment in young adulthood. Australian Orthodontic Journal, 21(2), 87–93.**  **Broadbent, J. M., Poulton, R., & Thomson, W. M. (2007). Trajectories of dental anxiety in a birth cohort. Journal of Dental Research, 86(6), 553–556. https://doi.org/10.1177/154405910708600613**  **Evans, R. W., Beck, D. J., & Brown, R. H. (1980). Dental health of 5-year-old children: a report from the Dunedin Multidisciplinary Child Development Study. New Zealand Dental Journal, 76(346), 179–186.**  **Poulton, R., Moffitt, T. E., & Silva, P. A. (2015). The Dunedin Multidisciplinary Health and Development Study: overview of the first 40 years, with an eye to the future. Social Psychiatry and Psychiatric Epidemiology, 50, 679–693. https://doi.org/10.1007/s00127-015-1048-8**  **Poulton, R., Thomson, W. M., Davies, S., Kruger, E., Brown, R. H., & Silva, P. (1997). Good teeth, bad teeth and fear of the dentist. Behaviour Research and Therapy, 35(4), 327–334. https://doi.org/10.1016/s0005-7967(96)00096-4**  **Poulton, R., Waldie, K. E., Craske, M. G., Menzies, R. G., & McGee, R. (2000). Dishabituation processes in height fear and dental fear: an indirect test of the non-associative model of fear acquisition. Behaviour Research and Therapy, 38(9), 909–919. https://doi.org/10.1016/s0005-7967(99)00115-4**  **Poulton, R., Waldie, K. E., Menzies, R. G., Craske, M. G., & Silva, P. A. (2001). Failure to overcome 'innate' fear: a developmental test of the non-associative model of fear acquisition. Behaviour Research and Therapy, 39(1), 29–43. https://doi.org/10.1016/s0005-7967(99)00153-1**  **Ruiz, B., Broadbent, J. M., Thomson, W. M., Ramrakha, S., Boden, J., Horwood, J., & Poulton, R. (2023). Is childhood oral health the ‘canary in the coal mine’ for poor adult general health? Findings from two New Zealand birth cohort studies. Community Dentistry and Oral Epidemiology, 51, 838–846. https://doi.org/10.1111/cdoe.12772**  **Thomson, W. M., Poulton, R., Broadbent, J. M., & Al-Kubaisy, S. (2006). Xerostomia and medications among 32-year-olds. Acta Odontologica Scandinavica, 64(4), 249–254.** [**https://doi.org/10.1080/00016350600582046**](https://doi.org/10.1080/00016350600582046)  **Evans, R. W., Beck, D. J., Silva, P. A., & Brown, R. H. (1982). Relationships between dental health behaviour and oral health status of 5-year-old children: a report from the Dunedin Multidisciplinary child development study. The New Zealand dental journal, 78(351), 11–16.**  **Beckley, A. L., Moffitt, T. E., & Poulton, R. (2021). The Dunedin Multidisciplinary Health and Development Study: Methods of a 40+ year longitudinal study. In J. C. Barnes (Ed.), The Encyclopedia of Research Methods in Criminology and Criminal Justice.** |
| 8 | **Environments for Healthy Living – the Griffith Birth Cohort study (EFHL)** | **Cameron, C. M., Scuffham, P. A., Spinks, A., Scott, R., Sipe, N., Ng, S. K., Wilson, A., Searle, J., Lyons, R. A., Kendall, E., Halford, K., Griffiths, L. R., Homel, R., & McClure, R. J. (2012). Environments for Healthy Living (EFHL) Griffith Birth Cohort Study: Background and methods. Maternal and Child Health Journal, 16, 1896–1905. https://doi.org/10.1007/s10995-011-0940-4**  **Fernando, S., Kumar, S., Bakr, M., Speicher, D., Lea, R., Scuffham, P. A., & Johnson, N. W. (2018). Children's untreated decay is positively associated with past caries experience and with current salivary loads of mutans Streptococci; negatively with self-reported maternal iron supplements during pregnancy: A multifactorial analysis. Journal of Public Health Dentistry, 78(4), 305–314. https://doi.org/10.1111/jphd.12282**  **Fernando, S., Speicher, D. J., Bakr, M. M., Benton, M. C., Lea, R. A., Scuffham, P. A., Mihala, G., & Johnson, N. W. (2015). Protocol for assessing maternal, environmental and epigenetic risk factors for dental caries in children. BMC Oral Health, 15, Article 167. https://doi.org/10.1186/s12903-015-0143-2**  **Fernando, S., Tadakamadla, S. K., Bakr, M., Scuffham, P. A., & Johnson, N. W. (2018). Indicators of risk for dental caries in children: A holistic approach. Pediatric Dentistry, 40(3), 207–211.** |
| 9 | **Gudaga Study** | **Comino, E., Craig, P., Harris, E., McDermott, D., Harris, M., Henry, R., Pulver, L. J., Kemp, L., & Knight, J. (2010). The Gudaga Study: establishing an Aboriginal birth cohort in an urban community. Australian and New Zealand Journal of Public Health, 34(S1), S9-S17. https://doi.org/10.1111/j.1753-6405.2010.00546.x**  **George, A., Grace, R., Elcombe, E., Villarosa, A. R., Mack, H. A., Kemp, L., Ajwani, S., Wright, D. C., Anderson, C., Bucknall, N., & Comino, E. (2018). The oral health behaviours and fluid consumption practices of young urban Aboriginal preschool children in south-western Sydney, New South Wales, Australia. Health Promotion Journal of Australia, 29(1), 24-31. https://doi.org/10.1002/hpja.29**  **George, A., Villarosa, A. R., Ingram, S., Fatema, K., Elliott, K., Grace, R., Kemp, L., Scharkie, S., Anderson, C., Bucknall, N., Wright, D. C., & Comino, E. (2021). Oral health status, behaviours, food and beverage consumption of aboriginal children in Australia. Health Promotion Journal of Australia, 32(S1), 113-122. https://doi.org/10.1002/hpja.354** |
| 10 | **Longitudinal Study of Australian Children (LSAC)** | **Kaunein, N., Singh, A., & King, T. (2020). Associations between Individual-level and Area-level social disadvantage and oral health behaviours in Australian adolescents. Australian Dental Journal, 65(4), 286–293. https://doi.org/10.1111/adj.12792**  **Kilpatrick, N. M., Neumann, A., Lucas, N., Chapman, J., & Nicholson, J. M. (2012). Oral health inequalities in a national sample of Australian children aged 2-3 and 6-7 years. Australian Dental Journal, 57(1), 38–44. https://doi.org/10.1111/j.1834-7819.2011.01644.x**  **Liu, T., Lingam, R., Lycett, K., Mensah, F. K., Muller, J., Hiscock, H., Huque, M. H., & Wake, M. (2018). Parent-reported prevalence and persistence of 19 common child health conditions. Archives of Disease in Childhood. https://doi.org/10.1136/archdischild-2017-313191**  **Stormon, N., Clifford, S., Lange, K., Mangoyana, C., Ford, P., Wake, M., & Lalloo, R. (2021). Oral health: Epidemiology and concordance in Australian children and parents. Community Dentistry and Oral Epidemiology, 49(6), 518-526. https://doi.org/10.1111/cdoe.12662**  **Stormon, N., Ford, P. J., & Lalloo, R. (2019). Oral health in the Longitudinal Study of Australian Children: An age, period, and cohort analysis. International Journal of Paediatric Dentistry, 29(4), 411-418. https://doi.org/10.1111/ipd.12485**  **Stormon, N., Ford, P. J., & Lalloo, R. (2020). Family-level predictors of Australian children's dental caries and injuries. Pediatric Dentistry, 42(1), 32–38.**  **Fleitas Alfonzo, L., Bentley, R., & Singh, A. (2021). Home ownership, income and oral health of children in Australia—A population-based study. Community Dentistry and Oral Epidemiology, 49(6), 543-550. https://doi.org/10.1111/cdoe.12646**  **Goldfeld, S., Francis, K. L., Hoq, M., Do, L., O'Connor, E., & Mensah, F. (2019). The impact of policy modifiable factors on inequalities in rates of child dental caries in Australia. International Journal of Environmental Research and Public Health, 16(11), 2029.**  **Hooley, M., Skouteris, H., & Millar, L. (2012). The relationship between childhood weight, dental caries and eating practices in children aged 4–8 years in Australia, 2004–2008. Pediatric Obesity, 7(6), 461-470. https://doi.org/10.1111/j.2047-6310.2012.00072.x**  **Lucas, N., Neumann, A., Kilpatrick, N., & Nicholson, J. M. (2011). State-level differences in the oral health of Australian preschool and early primary school-age children. Australian Dental Journal, 56(1), 56–62. https://doi.org/10.1111/j.1834-7819.2010.01287.x**  **Sawhney, S., Vu, T., Chen, F., Wong, K., Zafar, S., & Lopez Silva, C. P. (2023). Association between disability status and dental attendance patterns in Australian children: A national survey. Community Dentistry and Oral Epidemiology, 51(4), 746-753. https://doi.org/10.1111/cdoe.12755**  **Stormon, N., & Sexton, C. (2023). Parental recall bias in observational studies: Child dental service use. International Journal of Paediatric Dentistry, 33(4), 369-377. https://doi.org/10.1111/ipd.13051** |
| 11 | **Longitudinal Study of Indigenous Children (LSIC)** | **Toh, J. R., Wooi, N., Tan, S. N., Wong, K., Lopez-Silva, C., & Zafar, S. (2022). Association between lack of dental service utilisation and caregiver-reported caries in Australian Indigenous children: A national survey. Journal of Paediatrics and Child Health, 58(12), 2218–2224. https://doi.org/10.1111/jpc.16192**  **Islam, M. I., Chadwick, V., Esgin, T., & Martiniuk, A. (2022). Bullied Because of Their Teeth: Evidence from a Longitudinal Study on the Impact of Oral Health on Bullying Victimization among Australian Indigenous Children. International Journal of Environmental Research and Public Health, 19(9), 4995. https://doi.org/10.3390/ijerph19094995**  **Ju, X., Jamieson, L. M., & Mejia, G. C. (2016). Estimating the effects of maternal education on child dental caries using marginal structural models: The Longitudinal Study of Indigenous Australian Children. Community Dentistry and Oral Epidemiology, 44(5), 459-466. https://doi.org/10.1111/cdoe.12259**  **Claudia, C., Ju, X., Mejia, G., & Jamieson, L. (2016). The relationship between maternal smoking during pregnancy and parental-reported experience of dental caries in Indigenous Australian children. Community Dental Health, 33(4), 297–302. https://doi.org/10.1922/CDH_3937Claudia06**  **Thurber, K. A., Banks, E., & Banwell, C. (2015). Cohort Profile: Footprints in Time, the Australian Longitudinal Study of Indigenous Children. International Journal of Epidemiology, 44(3), 789–800. https://doi.org/10.1093/ije/dyu122** |
| 12 | **Pacific Islands Families Study (PIF)** | **Paterson, J. E., Gao, W., Sundborn, G., & Cartwright, S. (2011). Maternal self-report of oral health in six-year-old Pacific children from South Auckland, New Zealand. Community Dentistry and Oral Epidemiology, 39(1), 19–28. https://doi.org/10.1111/j.1600-0528.2010.00575.x**  **Paterson, J., Percival, T., Schluter, P., Sundborn, G., Abbott, M., Carter, S., Cowley-Malcolm, E., Borrows, J., Gao, W., & the PIF Study Group. (2008). Cohort Profile: The Pacific Islands Families (PIF) Study. International Journal of Epidemiology, 37(2), 273–279. https://doi.org/10.1093/ije/dym171**  **Schluter, P. J., Durward, C., Cartwright, S., & Paterson, J. (2007). Maternal Self-Report of Oral Health in 4-Year-Old Pacific Children from South Auckland, New Zealand: Findings from the Pacific Islands Families Study. Journal of Public Health Dentistry, 67(2), 69–77. https://doi.org/10.1111/j.0022-4006.2007.00014.x**  **Schluter, P. J., Kanagaratnam, S., Taylor, S., & Tautolo, E.-S. (2017). Acculturation and its impact on the oral health status of Pacific children in New Zealand: findings from the Pacific Islands Families study. Journal of Public Health Dentistry, 77(S1), 3–9. https://doi.org/10.1111/jphd.12202**  **Sundborn, G., Paterson, J., Jhagroo, U., Taylor, S., Iusitini, L., Tautolo, E.-S., Savila, F., Hirao, A., & Oliver, M. (2011). Cohort profile: A decade on and strong – The Pacific Islands Families Study. Pacific Health Dialog, 17(2), 9–21.** |
| 13 | **Queensland Birth Cohort Study** | **Harrison-Barry, L., Elsworthy, K., Pukallus, M., Leishman, S. J., Boocock, H., Walsh, L. J., & Seow, W. K. (2020). The Queensland Birth Cohort Study for Early Childhood Caries: Results at 7 Years. JDR Clinical & Translational Research, 6(2), 205-215. https://doi.org/10.1177/2380084420981882**  **Plonka, K. A., Pukallus, M. L., Barnett, A. G., Walsh, L. J., Holcombe, T. F., & Seow, W. K. (2012). A longitudinal study comparing mutans streptococci and lactobacilli colonisation in dentate children aged 6 to 24 months. Caries Research, 46(4), 385–393. https://doi.org/10.1159/000339089**  **Plonka, K. A., Pukallus, M., Barnett, A., Holcombe, T. F., Walsh, L. J., & Seow, W. K. (2012). A controlled, longitudinal study of home visits compared to telephone contacts to prevent early childhood caries. International Journal of Paediatric Dentistry, 23(1), 23-31. https://doi.org/10.1111/j.1365-263x.2011.01219.x**  **Plonka, K. A., Pukallus, M. L., Barnett, A. G., Holcombe, T. H., & Seow, W. K. (2013). A longitudinal case-control study of caries development from birth to 36 months. Caries Research, 47(2), 117–127. https://doi.org/10.1159/000345073**  **Plonka, K. A., Pukallus, M., Barnett, A. G., Walsh, L. J., Holcombe, T. H., & Seow, W. K. (2012). Mutans streptococci and lactobacilli colonization in predentate children from the neonatal period to seven months of age. Caries Research, 46(3), 213–220. https://doi.org/10.1159/000337353** |
| 14 | **South Australian Dental Longitudinal Study (SADLS)** | **Harrison-Barry, L., Elsworthy, K., Pukallus, M., Leishman, S. J., Boocock, H., Walsh, L. J., & Seow, W. K. (2020). The Queensland Birth Cohort Study for Early Childhood Caries: Results at 7 Years. JDR Clinical & Translational Research, 6(2), 205–215. https://doi.org/10.1177/2380084420981882**  **Ju, X., Harford, J., Luzzi, L., & Jamieson, L. M. (2022). Prevalence, extent, and severity of periodontitis among Australian older adults: Comparison of two generations. Journal of Periodontology, 93(9), 1387–1400. https://doi.org/10.1002/JPER.21-0458**  **Thomson, W. M., Chalmers, J. M., Spencer, A. J., & Slade, G. D. (2006). A longitudinal study of medication exposure and xerostomia among older people. Gerodontology, 23(4), 205–213. https://doi.org/10.1111/j.1741-2358.2006.00143.x**  **Thomson, W. M., Slade, G. D., Beck, J. D., Elter, J. R., Spencer, A. J., & Chalmers, J. M. (2004). Incidence of periodontal attachment loss over 5 years among older South Australians. Journal of Clinical Periodontology, 31(2), 119–125. https://doi.org/10.1111/j.0303-6979.2004.00454.x**  **Wang, M. T. M., Thomson, W. M., & Craig, J. P. (2019). Association between symptoms of xerostomia and dry eye in older people. Contact Lens and Anterior Eye, 42(6), 683-686.** [**https://doi.org/10.1016/j.clae.2019.09.002**](https://doi.org/10.1016/j.clae.2019.09.002)  **Hariyani, N., Spencer, A. J., Luzzi, L., & Do, L. G. (2018). Root surface caries among older Australians. Community Dentistry and Oral Epidemiology, 46(5), 484-493. https://doi.org/10.1111/cdoe.12386**  **Ju, X., Harford, J., Luzzi, L., Mejia, G., & Jamieson, L. M. (2022). A Longitudinal Study of Chronic Periodontitis in Two Cohorts of Community-Dwelling Elderly Australians. International Journal of Environmental Research and Public Health, 19(18), 11824. https://doi.org/10.3390/ijerph191811824**  **Slade, G. D., Gansky, S. A., & Spencer, A. J. (1997). Two-year incidence of tooth loss among South Australians aged 60+ years. Community Dentistry and Oral Epidemiology, 25(6), 429–437. https://doi.org/10.1111/j.1600-0528.1997.tb01734.x**  **Thomson, W. M., Chalmers, J. M., Spencer, A. J., & Slade, G. D. (2002). Is medication a risk factor for dental caries among older people? Evidence from a longitudinal study in South Australia. Community Dentistry and Oral Epidemiology, 30(3), 224–232. https://doi.org/10.1034/j.1600-0528.2002.300309.x**  **Thomson, W. M., Chalmers, J. M., Spencer, A. J., & Williams, S. M. (1999). The Xerostomia Inventory: A multi-item approach to measuring dry mouth. Community Dental Health, 16(1), 12–17.** |
| 15 | **Study of Mothers’ and Infants’ Life Events Affecting Oral Health (SMILE)** | **Bell, L. K., Schammer, C., Devenish, G., Ha, D., Thomson, M. W., Spencer, J. A., Do, L. G., Scott, J. A., & Golley, R. K. (2019). Dietary patterns and risk of obesity and early childhood caries in Australian toddlers: Findings from an Australian cohort study. Nutrients, 11(11), 2828. https://doi.org/10.3390/nu11112828**  **Devenish, G., Mukhtar, A., Begley, A., Spencer, A. J., Thomson, W. M., Ha, D., Do, L., & Scott, J. A. (2020). Early childhood feeding practices and dental caries among Australian preschoolers. The American Journal of Clinical Nutrition, 111(4), 821–828. https://doi.org/10.1093/ajcn/nqaa012**  **Do, L. G., Scott, J. A., Thomson, W. M., Stamm, J. W., Rugg-Gunn, A. J., Levy, S. M., Wong, C., Devenish, G., Ha, D. H., & Spencer, A. J. (2014). Common risk factor approach to address socioeconomic inequality in the oral health of preschool children - a prospective cohort study. BMC Public Health, 14(1), 429. https://doi.org/10.1186/1471-2458-14-429**  **Ha, D. H., & Do, L. G. (2018). Early life professional and layperson support reduce poor oral hygiene habits in toddlers—A prospective birth cohort study. Dentistry Journal, 6(4), 56. https://doi.org/10.3390/dj6040056**  **Ha, D. H., Spencer, A. J., Thomson, W. M., Scott, J. A., & Do, L. G. (2018). Commonality of risk factors for mothers’ poor oral health and general health: Baseline analysis of a population-based birth cohort study. Maternal and Child Health Journal, 22(5), 617–625.** [**https://doi.org/10.1007/s10995-018-2431-3**](https://doi.org/10.1007/s10995-018-2431-3)  **Do, L. G., Ha, D. H., Bell, L. K., Devenish, G., Golley, R. K., Leary, S. D., Manton, D. J., Thomson, W. M., Scott, J. A., & Spencer, A. J. (2020). Study of Mothers' and Infants' Life Events Affecting Oral Health (SMILE) birth cohort study: Cohort profile. BMJ Open, 10(10), e041185. https://doi.org/10.1136/bmjopen-2020-041185**  **Ha, D. H., Nguyen, H., Dao, A., Golley, R. K., Thomson, W. M., Manton, D. J., Leary, S. D., Scott, J. A., Spencer, A. J., & Do, L. G. (2022). Group-based trajectories of maternal intake of sugar-sweetened beverage and offspring oral health from a prospective birth cohort study. Journal of Dentistry, 122, 104113. https://doi.org/10.1016/j.jdent.2022.104113**  **Ha, D. H., Nguyen, H. V., Bell, L. K., Devenish-Coleman, G., Golley, R. K., Thomson, W. M., Manton, D. J., Leary, S. D., Scott, J. A., Spencer, J., & Do, L. G. (2023). Trajectories of child free sugars intake and dental caries – a population-based birth cohort study. Journal of Dentistry, 134, 104559. https://doi.org/10.1016/j.jdent.2023.104559**  **Hariyani, N., Do, L. G., Spencer, A. J., Thomson, W. M., Scott, J. A., & Ha, D. H. (2020). Maternal caries experience influences offspring’s early childhood caries—a birth cohort study. Community Dentistry and Oral Epidemiology, 48(6), 483-491. https://doi.org/10.1111/cdoe.12568** |
| 16 | **The 45 and Up Study** | **45 and Up Study Collaborators. (2008). Cohort profile: The 45 and Up Study. International Journal of Epidemiology, 37(5), 941–947. https://doi.org/10.1093/ije/dym184**  **Arora, M., Schwarz, E., Sivaneswaran, S., & Banks, E. (2010). Cigarette smoking and tooth loss in a cohort of older Australians: The 45 and Up Study. The Journal of the American Dental Association, 141(10), 1242–1249. https://doi.org/10.14219/jada.archive.2010.0354**  **Bleicher, K., Summerhayes, R., Baynes, S., Swarbrick, M., Navin Cristina, T., Luc, H., Dawson, G., Cowle, A., Dolja-Gore, X., & McNamara, M. (2023). Cohort Profile Update: The 45 and Up Study. International Journal of Epidemiology, 52(1), e92–e101. https://doi.org/10.1093/ije/dyac104**  **Gibson, A. A., Cox, E., Gale, J., Craig, M. E., Eberhard, J., King, S., Chow, C. K., Colagiuri, S., & Nassar, N. (2023). Oral health status and risk of incident diabetes: A prospective cohort study of 213,389 individuals aged 45 and over. Diabetes Research and Clinical Practice, 202, 110821. https://doi.org/10.1016/j.diabres.2023.110821**  **Gibson, A. A., Cox, E., Gale, J., Craig, M. E., King, S., Chow, C. K., Colagiuri, S., & Nassar, N. (2023). Association of oral health with risk of incident micro and macrovascular complications: A prospective cohort study of 24,862 people with diabetes. Diabetes Research and Clinical Practice, 203, 110857. https://doi.org/10.1016/j.diabres.2023.110857**  **Joshy, G., Arora, M., Korda, R. J., Chalmers, J., & Banks, E. (2016). Is poor oral health a risk marker for incident cardiovascular disease hospitalisation and all-cause mortality? Findings from 172 630 participants from the prospective 45 and Up Study. BMJ Open, 6(8), e012386. https://doi.org/10.1136/bmjopen-2016-012386** |
| 17 | **The Child Fluoride Study (CFS)** | **Slade, G. D., Davies, M. J., Spencer, A. J., & Stewart, J. F. (1995). Associations between exposure to fluoridated drinking water and dental caries experience among children in two Australian states. Journal of Public Health Dentistry, 55(4), 218–228. https://doi.org/10.1111/j.1752-7325.1995.tb02381.x**  **Slade, G. D., Spencer, A. J., Davies, M. J., & Burrow, D. (1996). Intra-oral distribution and impact of caries experience among South Australian school children. Australian Dental Journal, 41(5), 343–350. https://doi.org/10.1111/j.1834-7819.1996.tb03157.x**  **Spencer, A. J., & Do, L. G. (2008). Changing risk factors for fluorosis among South Australian children. Community Dentistry and Oral Epidemiology, 36(3), 210–218. https://doi.org/10.1111/j.1600-0528.2007.00389.x**  **Spencer, A. J., Armfield, J. M., & Slade, G. D. (2008). Exposure to water fluoridation and caries increment. Community Dental Health, 25(2), 68–73. https://doi.org/10.1922/CDH_2067Spencer11** |
| 18 | **Peri/Postnatal Epigenetic Twins Study (PETS)** | **Silva, M. J., Kilpatrick, N. M., Craig, J. M., Manton, D. J., Leong, P., Burgner, D., & Scurrah, K. J. (2018). Etiology of hypomineralized second primary molars: A prospective twin study. Journal of Dental Research, 97(13), 1441–1447. https://doi.org/10.1177/0022034518792870**  **Silva, M. J., Kilpatrick, N. M., Craig, J. M., Manton, D. J., Leong, P., Burgner, D. P., & Scurrah, K. J. (2019). Genetic and early-life environmental influences on dental caries risk: A twin study. Pediatrics, 143(5), e20183499. https://doi.org/10.1542/peds.2018-3499**  **Silva, M. J., Kilpatrick, N. M., Craig, J. M., Manton, D. J., Leong, P., Ho, H., Saffery, R., Burgner, D. P., & Scurrah, K. J. (2020). A twin study of body mass index and dental caries in childhood. Scientific Reports, 10(1), 568. https://doi.org/10.1038/s41598-020-57435-7** |
| 19 | **VicGeneration study (VicGen)** | **Carpenter, L., Gibbs, L., Magarey, A., Dashper, S., Gussy, M., & Calache, H. (2021). Nutrition and oral health in early childhood: associations with formal and informal childcare. Public Health Nutrition, 24(6), 1438–1448. https://doi.org/10.1017/S1368980020001676**  **Chattopadhyay, A., Christian, B., Masood, M., Calache, H., Carpenter, L., Gibbs, L., & Gussy, M. (2020). Natural history of dental caries: Baseline characteristics of the VicGen birth cohort study. International Journal of Paediatric Dentistry, 30(3), 334–341. https://doi.org/10.1111/ipd.12609**  **Christian, B., Calache, H., Adams, G., Hall, M., Dashper, S., Gibbs, L., & Gussy, M. (2020). A methodological study to assess the measurement properties (reliability and validity) of a caries risk assessment tool for young children. Journal of Dentistry, 95, 103324. https://doi.org/10.1016/j.jdent.2020.103324**  **Dashper, S. G., Mitchell, H. L., Lê Cao, K. A., Carpenter, L., Gussy, M. G., Calache, H., Gladman, S. L., Bulach, D. M., Hoffmann, B., Catmull, D. V., Pruilh, S., Johnson, S., Gibbs, L., Amezdroz, E., Bhatnagar, U., Seemann, T., Mnatzaganian, G., Manton, D. J., & Reynolds, E. C. (2019). Temporal development of the oral microbiome and prediction of early childhood caries. Scientific Reports, 9(1), 19732. https://doi.org/10.1038/s41598-019-56233-0**  **Gussy, M., Ashbolt, R., Carpenter, L., Virgo-Milton, M., Calache, H., Dashper, S., Leong, P., de Silva, A., de Livera, A., Simpson, J., & Waters, E. (2016). Natural history of dental caries in very young Australian children. International Journal of Paediatric Dentistry, 26(3), 173–183. https://doi.org/10.1111/ipd.12169**  **Gussy, M., Mnatzaganian, G., Dashper, S., Carpenter, L., Calache, H., Mitchell, H., Reynolds, E., Gibbs, L., Hegde, S., Adams, G., Johnson, S., Amezdroz, E., & Christian, B. (2020). Identifying predictors of early childhood caries among Australian children using sequential modelling: Findings from the VicGen birth cohort study. Journal of Dentistry, 93, 103276. https://doi.org/10.1016/j.jdent.2020.103276**  **Johnson, S., Carpenter, L., Amezdroz, E., Dashper, S., Gussy, M., Calache, H., de Silva, A. M., & Waters, E. (2017). Cohort Profile: The VicGeneration (VicGen) study: An Australian oral health birth cohort. International Journal of Epidemiology, 46(1), 29–30g. https://doi.org/10.1093/ije/dyw024** |
| 20 | **Growing Up in New Zealand** | **Thornley, S., Turton, B., Bach, K., Bird, A., Farrar, R., Bronte, S., Atatoa Carr, P., Fa'alili-Fidow, J., Morton, S., & Grant, C. (2021). What factors are associated with early childhood dental caries? A longitudinal study of the Growing Up in New Zealand cohort. International Journal of Paediatric Dentistry, 31(3), 351–360. https://doi.org/10.1111/ipd.12686** |
| 21 | **Healthy Smiles Healthy Kids (HSHK)** | **Arora, A., Scott, J. A., Bhole, S., Do, L., Schwarz, E., & Blinkhorn, A. S. (2011). Early childhood feeding practices and dental caries in preschool children: a multi-centre birth cohort study. BMC Public Health, 11(1), 28.** [**https://doi.org/10.1186/1471-2458-11-28**](https://doi.org/10.1186/1471-2458-11-28)  **Manohar, N., Hayen, A., Scott, J. A., Do, L. G., Bhole, S., & Arora, A. (2021). Impact of dietary trajectories on obesity and dental caries in preschool children: Findings from the Healthy Smiles Healthy Kids study. Nutrients, 13(7), 2240. https://doi.org/10.3390/nu13072240** |
| 22 | **South Australian Aboriginal Birth Cohort Study (SAABC)** | **Jamieson, L. M., Hedges, J., Ju, X., Kapellas, K., Leane, C., Haag, D. G., Santiago, P. R., Macedo, D. M., Roberts, R. M., & Smithers, L. G. (2021). Cohort profile: South Australian Aboriginal Birth Cohort (SAABC)—A prospective longitudinal birth cohort. BMJ Open, 11(3), e043559. https://doi.org/10.1136/bmjopen-2020-043559** |
| 23 | **Splash! Longitudinal Birth Cohort Study** | **de Silva-Sanigorski, A. M., Waters, E., Calache, H., Smith, M., Gold, L., Gussy, M., Scott, A., Lacy, K., & Virgo-Milton, M. (2011). Splash!: a prospective birth cohort study of the impact of environmental, social and family-level influences on child oral health and obesity related risk factors and outcomes. BMC Public Health, 11, 505. https://doi.org/10.1186/1471-2458-11-505**  **Martin-Kerry, J., Gussy, M., Gold, L., Calache, H., Boak, R., Smith, M., & de Silva, A. (2020). Are Australian parents following feeding guidelines that will reduce their child's risk of dental caries? Child: Care, Health and Development, 46(4), 495–505. https://doi.org/10.1111/cch.12768** |
| 24 | **Oral HPV in Indigenous South Australians cohort study** | **Jamieson, L. M., Garvey, G., Hedges, J., Leane, C., Hill, I., Brown, A., Ju, X., Sethi, S., Roder, D., Logan, R. M., Johnson, N., Smith, M., Antonsson, A., & Canfell, K. (2021). Cohort profile: indigenous human papillomavirus and oropharyngeal squamous cell carcinoma study - a prospective longitudinal cohort. BMJ Open, 11(6), e046928. https://doi.org/10.1136/bmjopen-2020-046928**  **Ju, X., Sethi, S., Antonsson, A., Hedges, J., Canfell, K., Smith, M., Garvey, G., Logan, R. M., & Jamieson, L. M. (2023). Natural History of Oral HPV Infection among Indigenous South Australians. Viruses, 15(7), 1573. https://doi.org/10.3390/v15071573** |
| 25 | **Dental Fluorosis Among South Australian Children** | **Do, L. G., & Spencer, A. J. (2007). Decline in the prevalence of dental fluorosis among South Australian children. Community Dentistry and Oral Epidemiology, 35(4), 282–291. https://doi.org/10.1111/j.1600-0528.2007.00314.x**  **Do, L. G., & Spencer, A. J. (2007). Oral health-related quality of life of children by dental caries and fluorosis experience. Journal of Public Health Dentistry, 67(3), 132–139. https://doi.org/10.1111/j.1752-7325.2007.00036.x**  **Do, L. G., & Spencer, A. J. (2007). Risk-benefit balance in the use of fluoride among young children. Journal of Dental Research, 86(8), 723–728.**  **Do, L. G., Ha, D. H., & Spencer, A. J. (2016). Natural history and long-term impact of dental fluorosis: A prospective cohort study. Medical Journal of Australia, 204(1), 25.e1. https://doi.org/10.5694/mja15.00703** |
| 26 | **Tooth Eruption in Infants Cohort Study** | **Wake, M., Hesketh, K., & Lucas, J. (2000). Teething and tooth eruption in infants: A cohort study. Pediatrics, 106(6), 1374–1379. https://doi.org/10.1542/peds.106.6.1374** |
| 27 | **Sugar–starch combinations in food and the relationship to dental caries in low-risk adolescents** | **Campain, A. C., Morgan, M. V., Evans, R. W., Ugoni, A., Adams, G. G., Conn, J. A., & Watson, M. J. (2003). Sugar-starch combinations in food and the relationship to dental caries in low-risk adolescents. European Journal of Oral Sciences, 111(4), 316–325.** |
| 28 | **Dental caries in Taranaki Adolescents Cohort Study** | **Foster Page, L. A., & Thomson, W. M. (2011). Dental caries in Taranaki adolescents: A cohort study. New Zealand Dental Journal, 107(3), 91–96.** |
| 29 | **The New Zealand Very Low Birth Weight Study (VLBW)** | **Darlow, B. A., Horwood, L. J., Woodward, L. J., Elliott, J. M., Troughton, R. W., Elder, M. J., Epton, M. J., Stanton, J. D., Swanney, M. P., Keenan, R., Melzer, T. R., McKelvey, V. A., Levin, K., Meeks, M. G., Espiner, E. A., Cameron, V. A., & Martin, J. (2015). The New Zealand 1986 very low birth weight cohort as young adults: Mapping the road ahead. BMC Pediatrics, 15, Article 90. https://doi.org/10.1186/s12887-015-0413-9**  **McKelvey, V., Darlow, B. A., Horwood, L. J., & Martin, J. (2021). Dental status of young adults born with very low birthweight: A national cohort study. Community Dentistry and Oral Epidemiology, 49(3), 240–248. https://doi.org/10.1111/cdoe.12595** |
| 30 | **Mater University of Queensland Study of Pregnancy (MUSP)** | **Bor, W., Najman, J. M., Andersen, M., Morrison, J., & Williams, G. (1993). Socioeconomic disadvantage and child morbidity: An Australian longitudinal study. Social Science & Medicine, 36(8), 1053–1061.** |
| 31 | **ASPREE Longitudinal Study of Older Persons (ALSOP)** | **Khan, S., Chen, Y., Crocombe, L., Ivey, E., Owen, A. J., McNeil, J. J., Woods, R. L., Wolfe, R., Freak-Poli, R., Britt, C., & Gasevic, D. (2024). Self-reported oral health status, edentulism and all-cause mortality risk in 12 809 Australian older adults: A prospective cohort study. Australian Dental Journal, 69(2), 82–92. https://doi.org/10.1111/adj.12987**  **McNeil, J. J., Woods, R. L., Ward, S. A., Britt, C. J., Lockery, J. E., Beilin, L. J., & Owen, A. J. (2019). Cohort Profile: The ASPREE Longitudinal Study of Older Persons (ALSOP). International Journal of Epidemiology, 48(4), 1048–1049h. https://doi.org/10.1093/ije/dyy279** |
| 32 | **Oral Health at Five Years Study** | **Sanders, A. E., & Slade, G. D. (2010). Apgar score and dental caries risk in the primary dentition of five year olds. Australian Dental Journal, 55(3), 260–267. https://doi.org/10.1111/j.1834-7819.2010.01232.x**  **Slade, G. D., Sanders, A. E., Bill, C. J., & Do, L. G. (2006). Risk factors for dental caries in the five-year-old South Australian population. Australian Dental Journal, 51(2), 130–139. https://doi.org/10.1111/j.1834-7819.2006.tb00416.x** |
| 33 | **Oral Health of Critically Ill Children Cohort Study** | **Ullman, A., Long, D., & Lewis, P. (2011). The oral health of critically ill children: An observational cohort study. Journal of Clinical Nursing, 20(21-22), 3070–3080. https://doi.org/10.1111/j.1365-2702.2011.03797.x** |
| 34 | **Malocclusion and Oral Health-Related Quality of Life Cohort Study** | **Healey, D. L., Gauld, R. D. C., & Thomson, W. M. (2016). Treatment-associated changes in malocclusion and oral health–related quality of life: A 4-year cohort study. American Journal of Orthodontics and Dentofacial Orthopedics, 150(5), 811–817. https://doi.org/10.1016/j.ajodo.2016.04.019** |
| 35 | **University of Adelaide longitudinal study of growth and development** | **Floyd, B., & Littleton, J. (2006). Linear enamel hypoplasia and growth in an Australian Aboriginal community: Not so small, but not so healthy either. Annals of Human Biology, 33(4), 424–443. https://doi.org/10.1080/03014460600748184**  **Littleton, J., & Townsend, G. C. (2005). Linear enamel hypoplasia and historical change in a central Australian community. Australian Dental Journal, 50(2), 101–107. https://doi.org/10.1111/j.1834-7819.2005.tb00348.x** |
| 36 | **North Brisbane Children Cohort Study** | **Butten, K., Johnson, N. W., Hall, K. K., Anderson, J., Toombs, M., O'Grady, K. F., & King, N. (2019). Risk factors for oral health in young, urban, Aboriginal and Torres Strait Islander children. Australian Dental Journal, 64(1), 72–81. https://doi.org/10.1111/adj.12662** |
| 37 | **The Household, Income and Labour Dynamics in Australia (HILDA) Survey** | **Lopez Silva, C. P., Singh, A., Calache, H., Derbi, H. A., & Borromeo, G. L. (2021). Association between disability status and dental attendance in Australia—A population-based study. Community Dentistry and Oral Epidemiology, 49(1), 33–39. https://doi.org/10.1111/cdoe.12571** |

**Independent Data Linkage Studies**

| # | Study Title | APA Citations |
| --- | --- | --- |
| 1D | Dental procedures in children with or without intellectual disability and autism spectrum disorder in a hospital setting | **Azimi, S., Wong, K., Lai, Y. Y. L., Bourke, J., Junaid, M., Jones, J., Pritchard, D., Calache, H., Winters, J., Slack-Smith, L., & Leonard, H. (2022). Dental procedures in children with or without intellectual disability and autism spectrum disorder in a hospital setting. Australian Dental Journal, 67(4), 328–339. https://doi.org/10.1111/adj.12927** |
| 2D | Early childhood caries sequelae and relapse rates in an Australian public dental hospital | **Tsai, C., Li, A., Brown, S., Deveridge, C., El Gana, R., Kucera, A., Irving, M., & Kumar, H. (2023). Early childhood caries sequelae and relapse rates in an Australian public dental hospital. International Journal of Paediatric Dentistry, 33(1), 1–11. https://doi.org/10.1111/ipd.12969** |
| 3D | Health effects of dental amalgam exposure: a retrospective cohort study | **Bates, M. N., Fawcett, J., Garrett, N., Cutress, T., & Kjellstrom, T. (2004). Health effects of dental amalgam exposure: a retrospective cohort study. International Journal of Epidemiology, 33(4), 894–902. https://doi.org/10.1093/ije/dyh164** |
| 4D | Sugar, dental caries and the incidence of acute rheumatic fever- a cohort study of Māori and Pacific children | **Thornley, S., Marshall, R. J., Bach, K., Koopu, P., Reynolds, G., Sundborn, G., & Le Shwe Sin, W. (2017). Sugar, dental caries and the incidence of acute rheumatic fever: a cohort study of Māori and Pacific children. Journal of Epidemiology and Community Health, 71(4), 364–370. https://doi.org/10.1136/jech-2016-208219** |
| 5D | Socio-demographic and familial factors associated with hospital admissions and repeat admission for dental caries in early childhood: A population-based study | **Chen, R., Schneuer, F. J., Irving, M. J., Chow, C. K., Kumar, H., Tsai, C., Sohn, W., Spallek, H., Bell, J., & Nassar, N. (2022). Socio-demographic and familial factors associated with hospital admissions and repeat admission for dental caries in early childhood: A population-based study. Community Dentistry and Oral Epidemiology, 50(6), 539–547. https://doi.org/10.1111/cdoe.12708** |
| 6D | Dental Hospital Admissions in the Children of Mothers with an Alcohol-Related Diagnosis: A Population-Based, Data-Linkage Study | **O'Leary, C. M., & Slack-Smith, L. M. (2013). Dental hospital admissions in the children of mothers with an alcohol-related diagnosis: A population-based, data-linkage study. The Journal of Pediatrics, 163(2), 515–520.e1. https://doi.org/10.1016/j.jpeds.2013.02.020** |
| 7D | Oral cavity cancer treatment outcomes in Western Australia | **Hendriks, T., Cardemil, F., & Sader, C. (2019). Oral cavity cancer treatment outcomes in Western Australia. Australian Journal of Otolaryngology, 2, 20. https://doi.org/10.21037/ajo.2019.06.01** |
| 8D | Patterns and predictors of dental hospitalizations in patients with acquired brain injury from pre-injury to acute and post-acute injury | **Azimi, S., Troeung, L., & Martini, A. (2023). Patterns and predictors of dental hospitalizations in patients with acquired brain injury from pre-injury to acute and post-acute injury. NeuroRehabilitation, 53(3), 309–321. https://doi.org/10.3233/NRE-230145** |
| 9D | Oral cavity squamous cell carcinoma survival by biopsy type: a cancer registry study | **Frydrych, A. M., Parsons, R., Threlfall, T., Austin, N., Davies, G. R., Booth, D., & Slack-Smith, L. M. (2010). Oral cavity squamous cell carcinoma survival by biopsy type: a cancer registry study. Australian Dental Journal, 55(4), 378–384. https://doi.org/10.1111/j.1834-7819.2010.01257.x** |
| 10D | Malignant transformation in oral lichen planus and lichenoid lesions: a 14-year longitudinal retrospective cohort study of 829 patients in New Zealand | **Guan, G., Mei, L., Polonowita, A., Hussaini, H., Seo, B., & Rich, A. M. (2020). Malignant transformation in oral lichen planus and lichenoid lesions: A 14-year longitudinal retrospective cohort study of 829 patients in New Zealand. Oral Surgery, Oral Medicine, Oral Pathology and Oral Radiology, 130(4), 411–418. https://doi.org/10.1016/j.oooo.2020.07.002** |
